# Alcohol use disorder and body mass index show genetic pleiotropy and shared neural associations

**DOI:** 10.1101/2024.05.03.24306773

**Authors:** Samantha G. Malone, Christal N. Davis, Zachary Piserchia, Michael R. Setzer, Sylvanus Toikumo, Hang Zhou, Emma L. Winterlind, Joel Gelernter, Amy Justice, Lorenzo Leggio, Christopher T. Rentsch, Henry R. Kranzler, Joshua C. Gray

## Abstract

Despite neurobiological overlap, alcohol use disorder (AUD) and body mass index (BMI) show minimal genetic correlation (r_g_), possibly due to mixed directions of shared variants. We applied MiXeR to investigate shared genetic architecture between AUD and BMI, conjunctional false discovery rate (conjFDR) to detect shared loci and their directional effect, Local Analysis of (co)Variant Association (LAVA) for local r_g_, Functional Mapping and Annotation (FUMA) to identify lead single nucleotide polymorphisms (SNPs), Genotype-Tissue Expression (GTEx) to examine tissue enrichment, and BrainXcan to assess associations with brain phenotypes. MiXeR indicated 82.2% polygenic overlap, despite a r_g_ of −.03. ConjFDR identified 132 shared lead SNPs, with 53 novel, showing both concordant and discordant effects. GTEx analyses identified overexpression in multiple brain regions. Amygdala and caudate nucleus volumes were associated with AUD and BMI. Opposing variant effects explain the minimal rg between AUD and BMI, with implicated brain regions involved in executive function and reward, clarifying their polygenic overlap and neurobiological mechanisms.

## Introduction

Alcohol use disorder (AUD) and obesity adversely impact millions of individuals and contribute to hundreds of billions of dollars in combined annual economic cost^1,2^. The detrimental health impacts of AUD include increased risk for cancers of the liver, head, and neck; cardiovascular disease; and liver disease including cirrhosis^3^. Obesity, typically defined as a body mass index (BMI) of greater than 30 kg/m^21^, is associated with increased risk for hypertension, type II diabetes, coronary artery disease, liver disease, and various cancers^4^. The weighted prevalence of co-occurring heavy alcohol consumption and obesity in the United States rose from 1.8% in 1999-2000 to 3.1% in 2017-2020, representing a 72% increase^5^.

Alcohol use disorder is thought to share pathophysiological mechanisms with unhealthy eating behavior, a primary risk factor for elevated BMI and obesity (for additional causes of excess weight, see^6^). For example, the neurotransmitter dopamine plays integral roles in both eating and alcohol-related behaviors by impacting motivation, self-regulation, and reinforcement^7–9^. Evidence of other overlapping neurocircuitry-based mechanisms that contribute to AUD and pathological overeating has also accumulated^7,10^, which supports the concept of food addiction^11^, though this remains controversial^12^. There is also overlap in the interaction of both alcohol and food with appetite-related neuroendocrine pathways such as ghrelin ^13,14^ and glucagon-like peptide-1 (GLP-1)^15^. Interest in this putative overlap has been heightened by recent reports that GLP-1 receptor agonists (GLP-1RAs)—approved for treating type 2 diabetes and obesity—may represent potential new pharmacotherapies for AUD^16^. Similarly, medications used to treat AUD reduce weight in individuals with obesity^17^. For example, topiramate, recommended as an off-label treatment for AUD^18,19^, is approved in combination with phentermine as a weight loss medication.

Both AUD and obesity have substantial genetic contributions, with an estimated heritability of 50% for AUD ^20^ and 40-70% for obesity^21^. Despite shared neurobiological pathways and a high rate of co-occurrence in some populations, the genetic correlation between alcohol- and BMI-related phenotypes is non-significant^22,23^. In the two largest studies to date, null genetic correlations were reported between obesity and problematic alcohol use (i.e., a phenotype that combines AUD diagnoses and a quantitative measure of harmful drinking; r_g_ = −0.03) and drinks per week (r_g_ = 0.03) in European-ancestry individuals^22,23^.

Although these findings are consistent with a modest amount of shared genetic variation between the two traits, an alternative hypothesis is that the presence of shared variants with both concordant and discordant effects across the two phenotypes obscures evidence of genome-wide correlation. Other analytic methods, such as bivariate causal mixture models, are not influenced by the directionality of the variants’ effects, making them more appropriate to evaluate the extent of polygenic overlap between the two conditions^24^. For example, accounting for concordant and discordant variant effects has revealed substantial shared genetic relationships between psychiatric and medical or cognitive traits despite small or null genetic correlations^25^.

In this study, we utilized MiXeR to investigate the overall shared genomic architecture between AUD and BMI. MiXeR is a statistical method that estimates the potential causal variants for each trait and the total degree of overlap between two traits without regard to the direction of variants’ effects, thereby identifying the extent of unique and shared genetic architecture^24^. MiXeR has been used to identify genetic overlap across multiple psychiatric disorders^25^, psychiatric disorders and irritable bowel syndrome ^26^, and psychiatric disorders and cognitive traits such as educational attainment^27,28^. To complement MiXeR’s overarching approach, we also utilized Local Analysis of (co)Variant Association (LAVA) to estimate local regional genetic correlations^29^ and the conjunctional false discovery rate (conjFDR) method to identify specific overlapping loci^30^.We hypothesized that the absence of genetic correlation between AUD and BMI is attributable to the presence of shared variants with inconsistent directions of effect and that this would be evidenced by both (1) greater genetic overlap than would be predicted by the observed genetic correlation and (2) shared loci showing a mixture of consistent and inconsistent effect directions. After testing those hypotheses, we conducted follow-up functional annotation and drug repurposing analyses on shared loci identified by conjFDR. Additionally, we examined the associations of AUD and BMI risk with brain image-derived phenotypes (IDPs) to identify potential shared neural underpinnings. In doing so, we aimed to uncover the genomic architecture and physiologic pathways shared by AUD and BMI to advance our understanding of mechanisms contributing to their comorbidity and aid in the development of physiologically-informed interventions.

## Methods

### Samples

We used summary statistics from two large-scale GWAS for AUD (*N* = 753,248; *N*_case_ = 113,325)^23^ and BMI (*N* = 681,275)^31^. The AUD GWAS summary statistics were derived from a meta-analysis of several cohorts of individuals with AUD or alcohol dependence (AD) diagnoses and controls with no diagnosis. Despite differences in the specific criteria used for diagnosis across cohorts, the genetic correlation across diagnostic categories and cohorts was very high (Million Veteran Program AUD and Psychiatric Genomics Consortium AD r_g_ = .98)^32^. We excluded UK Biobank participants from the AUD GWAS to minimize the overlap of participants between the two studies and inflation by cross-trait enrichment^30^. The BMI GWAS summary statistics were derived from a meta-analysis of two cohorts: the Genetic Investigation of Anthropometric Traits (GIANT) consortium and the UK Biobank. All participants were of European ancestry (for more details, see original publications and Table S1). The Uniformed Services University’s Human Research Protections Program Office determined the project to be considered research not involving human subjects per 32 CFR 219.102(e)(1), and applicable DoD policy guidance.

### Characterizing polygenic overlap

MiXeR^24,33^ was applied to investigate the overall shared genetic architecture between AUD and BMI. Univariate MiXeR analyses were first conducted to estimate each trait’s polygenicity (i.e., the number of potential causal variants required to explain 90% of single nucleotide polymorphism [SNP] heritability) and discoverability (i.e., the average estimated effect size of causal variants). Prior to performing bivariate models, we confirmed that the Akaike and Bayesian information criteria (AIC and BIC) values of the univariate models were positive, supporting sufficient power for bivariate MiXeR analyses. Next, bivariate models were implemented to identify the number of unique and shared causal variants for each pair of traits. These models also provide estimates of the proportion of causal variants with concordant directions of effect. The Dice coefficient, an indicator of the proportion of polygenic overlap, is also computed. Conditional Q-Q plots were produced to visualize cross-trait enrichment. These plots show the distribution of p-values for a primary phenotype as a function of its association with the secondary phenotype at three p-value strata *(p* ≤ 0.1, 0.01, and 0.001). As a secondary analysis, we used MiXeR to examine the shared genetic architectures of AUD and BMI with other psychiatric traits (major depressive disorder, attention-deficit/hyperactivity disorder [ADHD], schizophrenia)^27,31,34,35^. This secondary analysis shows how the shared genetic architecture of AUD and BMI compares with that of other psychiatric traits. Finally, the shared genetic architecture of AUD and BMI with left-handedness was included as a control^36^. For all traits, LD Score regression (LDSC) v1.0.1 was used to calculate heritability^37^, genetic correlations, and standard errors.

Local Analysis of (co)Variant Association (LAVA) was used to estimate local heritability (*h*^2^_SNP_) and genetic correlations (r_g_) between AUD and BMI across 2,495 approximately equal-sized linkage disequilibrium (LD) blocks^29^. Significance of local *r*_g_ was adjusted using false discovery rate (FDR) correction^39^.

We used conjFDR^30,38^ analysis to detect loci significantly associated with both phenotypes, including variants with opposite directions of effect. To achieve this, conditional false discovery rate (condFDR) estimates were first obtained by conditioning the primary phenotype’s (i.e., AUD) test statistics on a secondary phenotype’s (i.e., BMI’s) SNP associations. A condFDR value for the second phenotype conditioned on the first’s SNP associations was calculated by reversing the order of phenotypes from the first condFDR assessment. ConjFDR defines the value for each association as the maximum of the two condFDR values for the given SNP, providing a conservative estimate of the SNP association with both phenotypes. Statistical significance was defined as a conjFDR value < 0.05. We performed a replication analysis of the significant loci from the conjFDR using summary statistics from FinnGen v11. Specifically, we utilized the phenotypes: “body-mass index, inverse-rank normalized” (*N* = 321,672) and “alcohol use disorder, Swedish definition” (*N* = 453,733; *N*_case_ = 26,149)^39^. Exact binomial tests (i.e., SNP sign tests) were performed to assess whether the shared genomic loci were collectively replicated in the independent samples^40^. The SNP sign test evaluates whether the directionality of allelic associations between the discovery and replication cohorts is consistent, with the null hypothesis being that the consistency of effect directions is 50%.

### Genomic loci definition and gene-set enrichment

SNPs having a conjFDR<.05, indicating significant SNP effects on both AUD and BMI, were input into Functional Mapping and Annotation (FUMA) v1.5.2^41^ to identify LD-independent genomic loci. Independent significant SNPs were identified using a LD block distance for merging of ≤ 250 kb, r^2^ < 0.6, and the European ancestry 1000 Genomes reference panel^42^. Of the independent SNPs, lead SNPs were identified using r^2^ < 0.1. Each locus is represented by a single lead SNP with the lowest conjFDR value. The novelty of lead SNPs was determined by examining whether variants were genome-wide significant (*p* < 5×10^-8^) in the AUD and BMI summary statistics. Lead SNPs were assigned to genes based on presence within the gene or otherwise distance to the nearest gene transcription start site. Annotations for the lead SNPs corresponding to Variant Effect Predictor (VEP), Combined Annotation Dependent Deletion (CADD) scores, and nearest transcription start site were sourced from OpenTargets (https://genetics.opentargets.org/ v22.10)^43^. The presence of lead SNPs within genes was confirmed using dbGaP (https://www.ncbi.nlm.nih.gov/gap/). Genes linked to the lead SNPs were then used to conduct gene expression, tissue enrichment specificity, and gene-set enrichment analyses in FUMA^41^. Gene expression analyses used transcripts per million (TPM) normalization, which is robust to differences in library size and sequencing depth across samples. Analyses were corrected for multiple comparisons using the FDR correction.

### Drug repurposing

We integrated drug-protein interaction/druggability information from the Target Central Resource Database (TCRD)^44^ and OpenTargets^43^. We searched for each gene in each database (https://pharos.nih.gov/ v3.18.0; https://platform.opentargets.org/ v23.12). The TCRD divides target development/druggability into four levels: (1) Tclin targets have approved drugs with known mechanisms of action; (2) Tchem targets have drugs or small molecules that satisfy activity thresholds; (3) Tbio targets have no known drugs or small molecules that satisfy thresholds, but have Gene Ontology (GO) leaf term annotations or Online Mendelian Inheritance in Man (OMIM) phenotypes, or meet at least two of three conditions: a fractional PubMed count >5, >3 National Center for Biotechnology Gene Reference Intro Function annotations, or >50 commercial antibodies; and (4) Tdark targets—proteins that have been manually curated in UniProt but do not meet criteria for the above categories. OpenTargets was utilized to identify approved drugs and drugs in development that target identified druggable genes.

### BrainXcan

We used BrainXcan software^45^ to evaluate the associations of AUD and BMI with 327 brain image-derived phenotypes (IDPs) obtained from structural (T1-weighted) and diffusion magnetic resonance imaging (dMRIs). BrainXcan infers trait-IDP associations using GWAS summary statistics, brain feature prediction weights, and reference LD data. Prediction weights for BrainXcan were derived by training a ridge regression model on brain IDPs in 24,409 individuals from the UK Biobank. Effect sizes and p-values of trait-IDP associations were adjusted using LD block-based permutation, and Bonferroni correction was used to account for multiple testing (T1: 0.05/109 = 4.59 × 10^-4^; dMRI = 0.05/218 = 2.29 × 10^-4^). Based on these results, we identified brain IDPs that were associated with both AUD and BMI. We also examined the concordance of effect direction for brain IDPs across the two traits.

## Results

### Shared global and local genomic architecture (MiXeR and LAVA)

All univariate AIC and BIC estimates were positive, with the exception of the more stringent BIC criterion for left-handedness (AIC = 3.62, BIC = −5.99), which indicated there was sufficient power overall for conducting bivariate MiXeR models. Bivariate AUD and BMI models show that the MiXeR estimated model had improved fit compared to the minimum overlap predicted by the genetic correlation alone (best vs. minimum AIC = 91.11, best vs. minimum BIC = 82.13, where positive values indicate improved fit) and fit significantly better than the maximum overlap model, which assumes maximum overlap of causal variants, when using AIC but not when considering the more stringent BIC (best vs. maximum AIC = 4.80, best vs. maximum BIC = −4.17); see Tables S2 and S3 for full univariate and bivariate MiXeR results).

MiXeR analysis yielded an overall level of polygenic overlap between AUD and BMI of 82.2% (as quantified by the Dice coefficient) despite a minimal genetic correlation (r_g_ = −0.03, SE = .02). Of the 9.4K and 11.1K potential causal variants linked to AUD and BMI, respectively, 8.4K were shared by the two traits (Figure 1). The estimated proportion of shared variants with a concordant direction of effect was 48.8%. Conditional Q-Q plots demonstrated enrichment of SNP associations with AUD that increased with the significance of the associations with BMI, and vice versa for BMI, both reflecting polygenic overlap (Figure S1).

**Figure 1.**
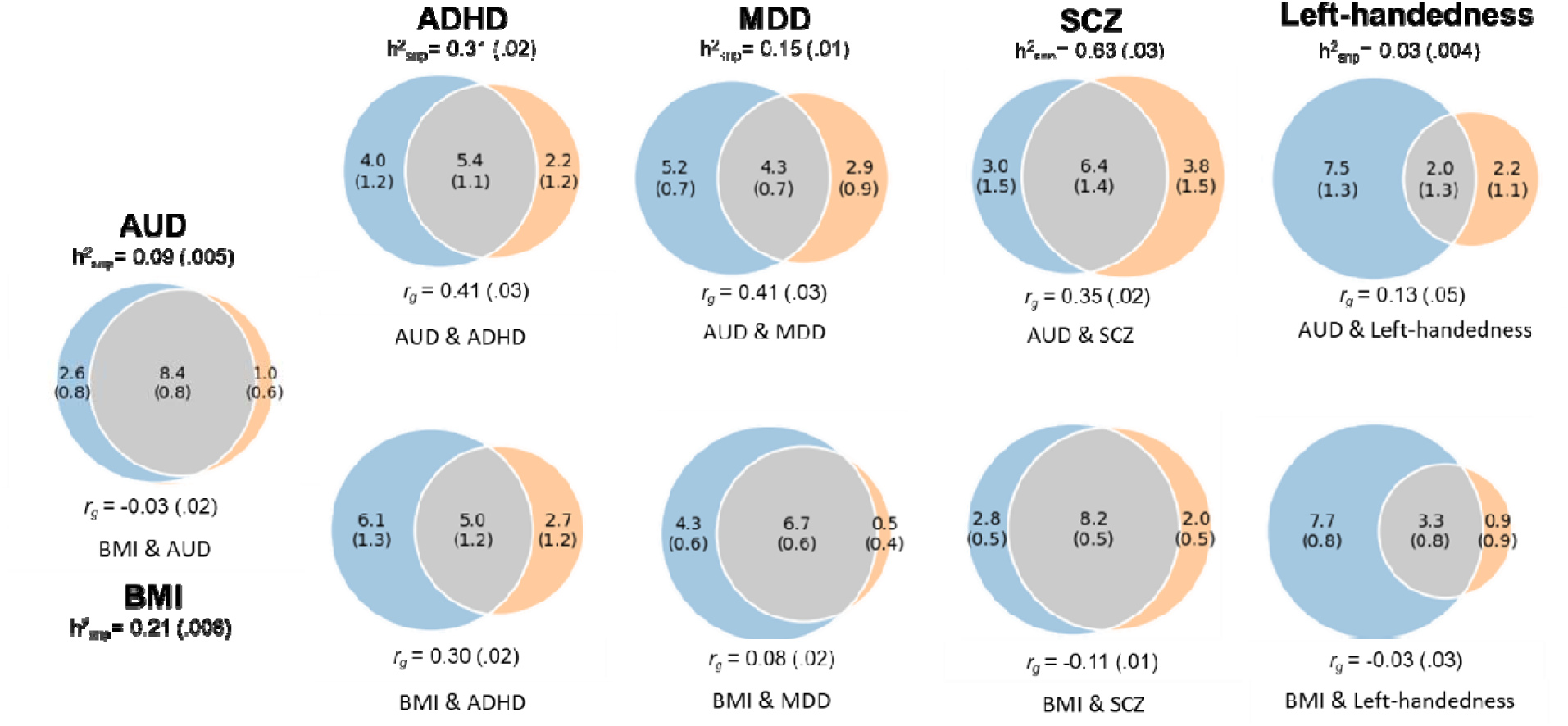
MiXeR Venn diagrams showing the estimated number of shared causal variants in the thousands and genetic correlations (*r_g_*) of AUD and BMI with each other and psychiatric disorders. Left-handedness was included as a control. h^2^_SNP_ = SNP-based heritability. Standard errors for genetic correlations and heritability estimates are included in parentheses. ADHD = attention-deficit/hyperactivity disorder, AUD = alcohol use disorder, BMI = body mass index, MDD = major depressive disorder, SCZ = schizophrenia.

The *r_g_*s between BMI and other psychiatric phenotypes ranged from −0.11 for schizophrenia to 0.30 for ADHD. There were substantial shared genetic variants between BMI and other traits, including 8.2K of the 10.2K schizophrenia variants (Dice coefficient = 77.3%; concordance = 45.0%), 6.7K of the 7.2K MDD variants (Dice coefficient = 73.3%; concordance = 54.6%), and 5.0K of the 7.7K ADHD variants (Dice coefficient = 53.3%; concordance = 74.8%). Finally, of the 4.2K potential causal variants linked to the control condition left-handedness, 3.3K (Dice coefficient = 43.4%; concordance = 46.8%) were shared with BMI. Of note, BMI had 7.7K potential causal variants that were not shared with left-handedness.

There were moderate genetic correlations (*r_g_*s = .35-.41) and shared genetic variants between AUD and other psychiatric traits, including 6.4K of the 10.2K schizophrenia variants (Dice coefficient = 65.6%; concordance = 71.9%), 4.3K of the 7.2K MDD variants (Dice coefficient = 51.4%; concordance = 90.8%), and 5.4K of the 7.7K ADHD variants (Dice coefficient = 63.5%; concordance = 74.8%). Finally, of the 4.2K potential causal variants linked to the control condition left-handedness, 2.0K (Dice coefficient = 28.3%; concordance = 73.2%) were shared with AUD (Figure 1).

Of the 2,495 independent regions examined in LAVA, local *h*^2^_SNP_ was significant (*p* < .05) in 2,293 regions for both AUD and BMI. Furthermore, local *r*_g_ between AUD and BMI was significant in 41 regions after FDR correction (*q* < .05). Just under half of these regions (19/41) had positive genetic correlations between the two phenotypes (Table S5).

### Shared genetic loci (cond/conj FDR)

At conjFDR<0.05, we identified 132 significant loci associated with both AUD and BMI (Figure 2). Most (121) of these variants were not significant in the original AUD GWAS and 53 were not significant in the original BMI GWAS. Notably, all 53 loci that were novel for BMI were also novel for AUD (Table S4). Of the shared loci, 56 lead SNPs (42.4%) had consistent effect directions for AUD and BMI and 76 (57.6%) had opposite effect directions. This supports the hypothesis that the lack of genetic correlation is due to mixed effect directions of variants shared by AUD and BMI.

**Figure 2.**
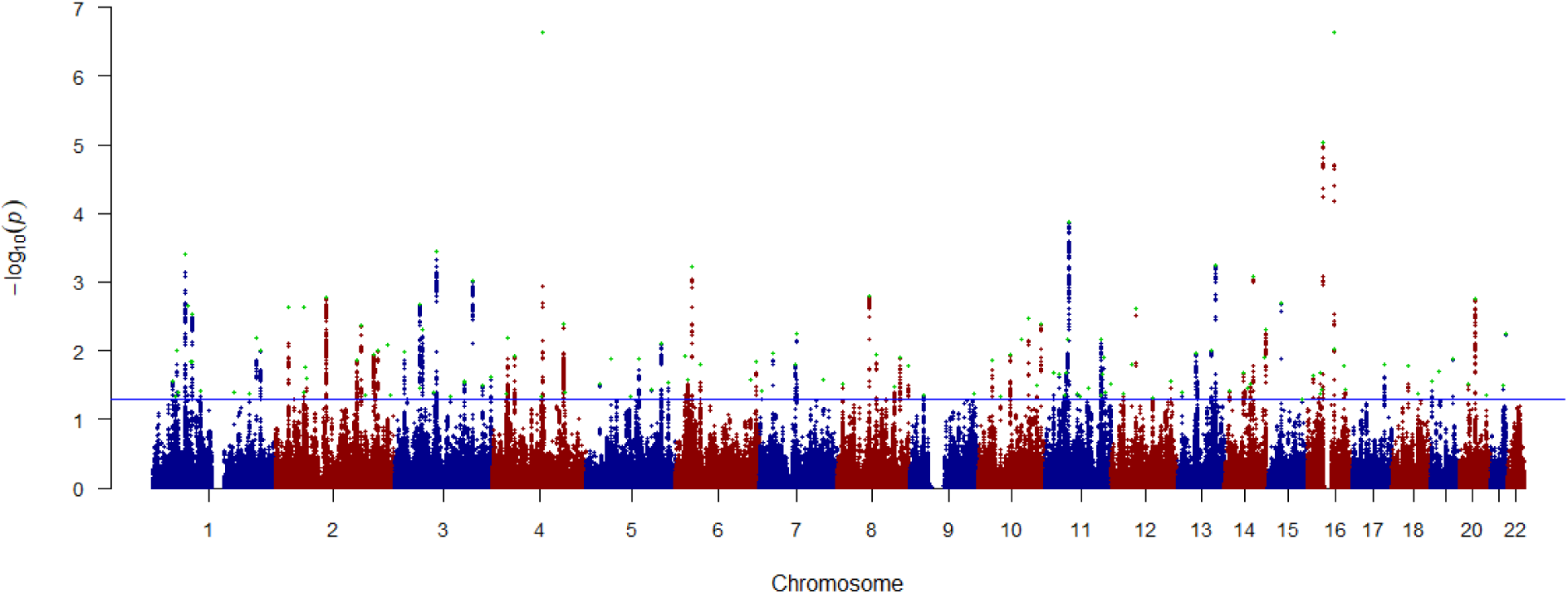
Manhattan plot of variants jointly associated with AUD and BMI. The plot depicts log_10_ transformed conjFDR values for each SNP on the y-axis and chromosomal position on the x-axis. The horizontal line is the threshold for significant shared associations between AUD and BMI (conjFDR < 0.05). Independent lead SNPs are green. The results are also shown in Table S4.

The two most significant loci exerted opposite directions of effect for BMI and AUD. The first was an intronic variant (rs9939973) in the well-known obesity risk gene, *FTO* (also linked to numerous other traits, including substance use disorders, major depression, and schizophrenia^46^. The second was an intronic variant (rs13114738) in the highly pleiotropic gene *SLC39A8*^47^. The most significant locus (rs10511087) with a concordant direction of effect was *CADM2*, which encodes cell adhesion molecule 2 and has been linked to array of cognition, pain, substance use, and metabolic phenotypes^48,49^.

Of the 2,495 independent regions examined in LAVA, local *h*^2^_SNP_ was significant (*p* < .05) in 2,293 regions for both AUD and BMI. Furthermore, local *r*_g_ between AUD and BMI was significant in 41 regions after FDR correction (*q* < .05). Just under half of these regions (19/41) had positive genetic correlations between the two phenotypes (Table S5).

### Replication in FinnGen

We performed replication analysis of the 132 significant loci using independent AUD (*N* = 453,733; *N*_case_ = 26,149) and BMI summary statistics (*N* = 321,672) from FinnGen v11. In the independent AUD summary statistics, 124 loci (94%) had sign concordance, 75 (57%) were nominally significant, and 55 (42%) were significant after FDR correction. In the independent BMI summary statistics, 125 loci (95%) had sign concordance, 97 (73%) were nominally significant, and 95 (72%) were significant after FDR correction (Table S4). The SNP sign tests supported the overall replication of loci effect directions across both the AUD (*p* < 2.2×10^-16^) and BMI (*p* < 2.2×10^-16^) independent cohorts. An important caveat to these analyses is that FinnGen sample is from an isolated population, and the AUD replication sample had only a quarter of the number of cases as the original discovery sample. The fact that a larger proportion of BMI loci replicated is likely attributable to more comparable sample sizes between the original and replication datasets (321,672/681,275 = 47%). The consistency of replication reported here is comparable to that found in other GWAS of complex traits^50–52^.

### Functional annotations

The Variant Effect Predictor (VEP) analysis of the 132 lead SNPs in the loci shared between AUD and BMI indicated that 74 were intronic, 31 intergenic, 7 downstream, 6 non coding transcript exon variant, 5 missense, 4 upstream, 3 in the 3’UTR, 1 synonymous, and 1 regulatory (Table S4). Ten of the 132 variants had CADD scores > 12.37, indicating possible deleteriousness^53^.

Analysis of Genotype-tissue Expression (GTEx)^54^ samples showed that genes linked to the lead SNPs were significantly overexpressed in the brain (Figure S2). Specifically, 47 genes were significantly upregulated in the frontal cortex (BA9), hypothalamus, cortex, anterior cingulate cortex (BA24), hippocampus, and amygdala (Figures 3 and 4). Analysis of GTEx samples for the shared genes linked to concordant loci identified significant upregulation in the hypothalamus and amygdala (Figure S4). Finally, analysis of the of the shared genes linked to discordant loci identified significant upregulation in the frontal cortex (BA9), hypothalamus, cortex, anterior cingulate cortex (BA24) (Figure S5).

**Figure 3.**
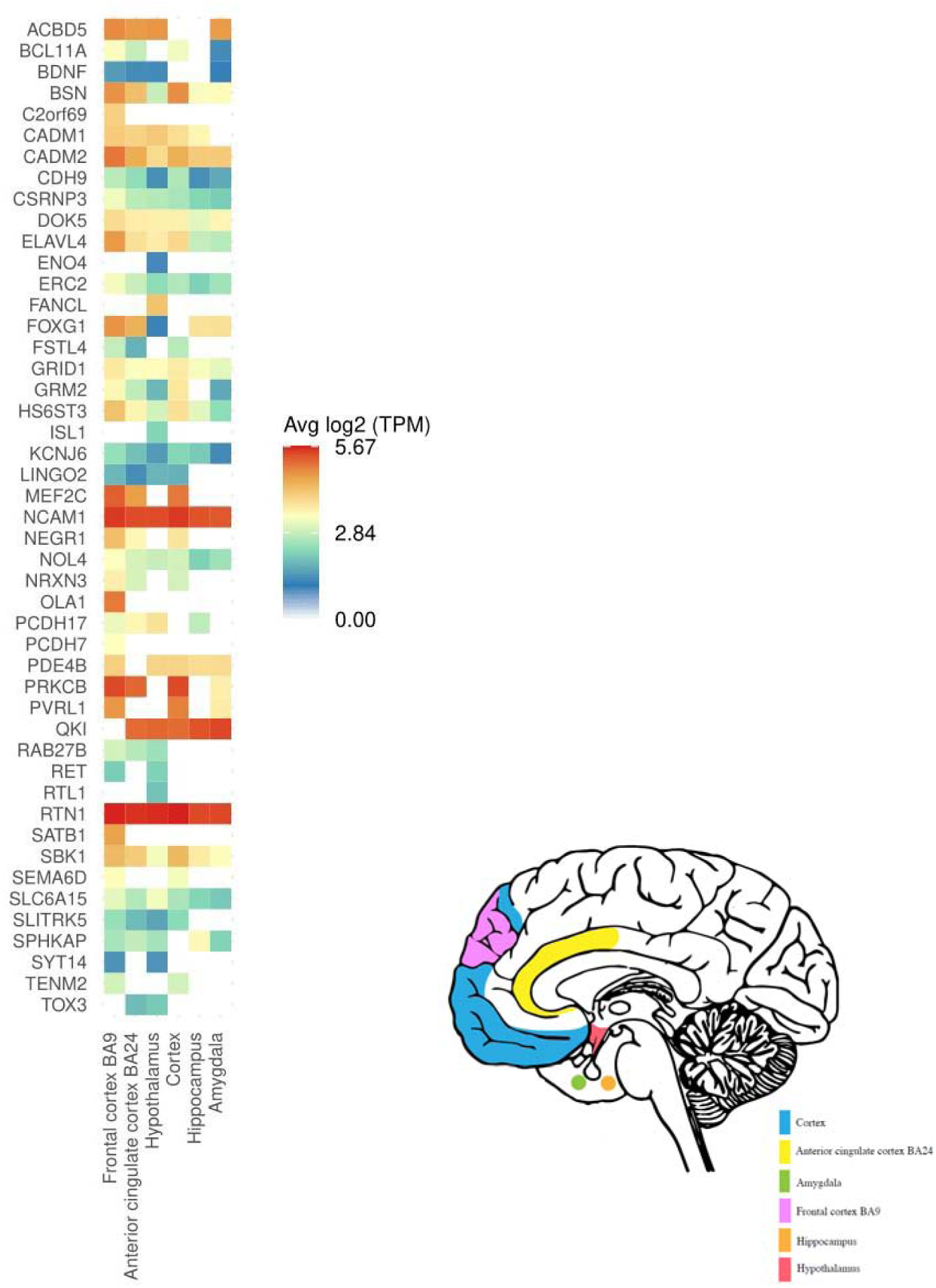
Significant up-regulated differential gene expression (DEG) in the frontal cortex (BA9), hypothalamus, cortex, anterior cingulate cortex (BA24), hippocampus, and amygdala. Tissues are significantly enriched at Bonferroni corrected p-value ≤ 0.05 for 54 GTEx tissue types. Only genes with a Bonferroni corrected p-value and absolute log fold change ≥ 0.58 for a given tissue are included. For full results across all brain tissues, regardless of significance, see Figure S3.

**Figure 4.**
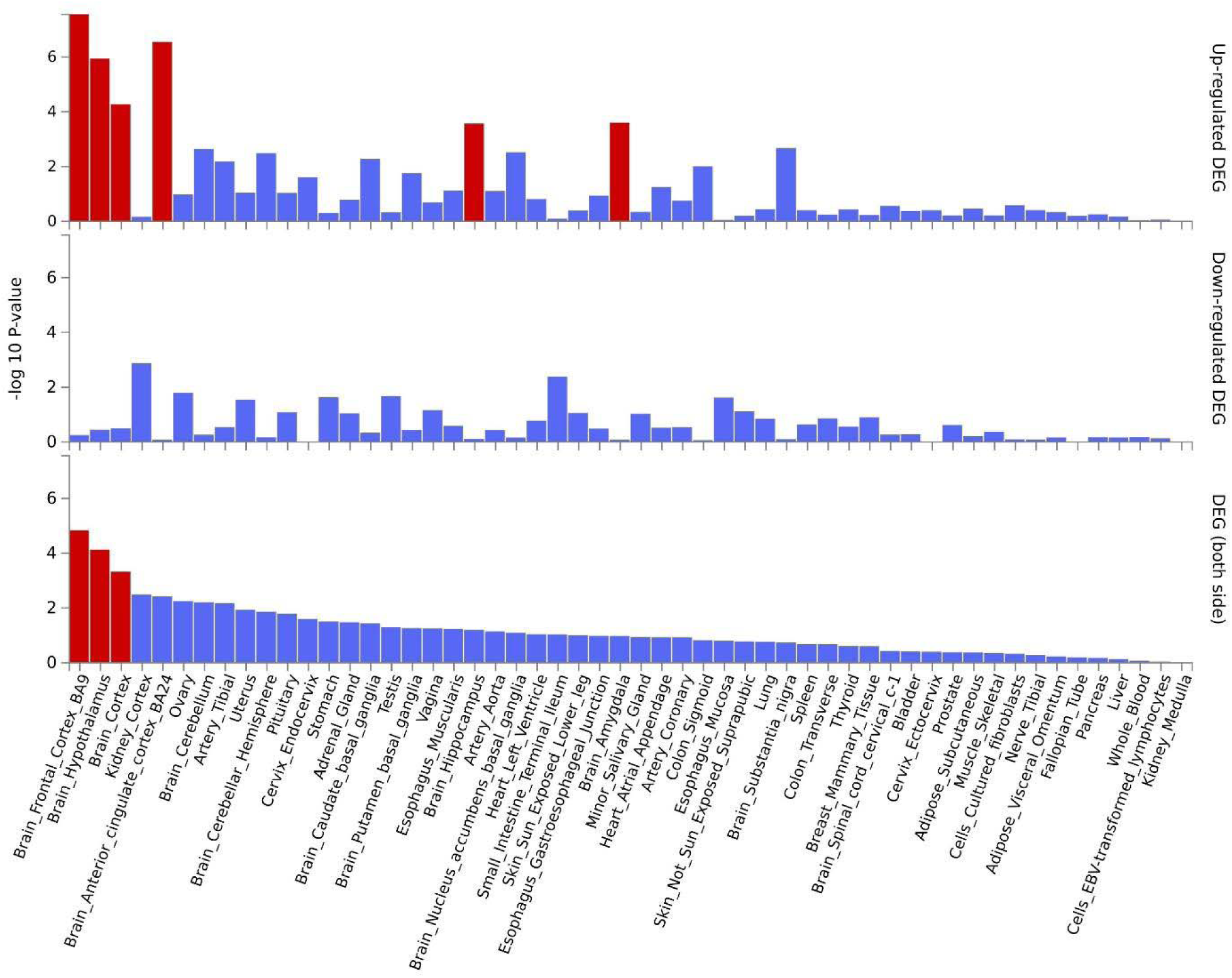
Differential gene expression (DEG) in 54 GTEx tissue types for genes linked to lead SNPs in distinct loci significantly associated with AUD and BMI. Significant enrichment (*p* < .05 after Bonferroni correction) is highlighted in red.

GO gene-set analysis identified 30 biological processes where shared genes were significantly enriched, with “cell morphogenesis involved in differentiation”, “cell morphogenesis”, “axon development”, and “presynaptic active zone organization” being the top four processes (Table S6). Additionally, genes were enriched for seven GO cellular component processes, including “synapse”, “GABAergic synapse”, and “presynapse”. Finally, the genes were enriched for 21 cell type signatures^55^, the top seven of which were in the midbrain and included “HGABA”, “HNBGABA”, “HDA1”, “HDA”, “HDA2”, “HNBML5”, and “HSERT”. These cell types are GABAergic, dopaminergic, serotonergic, and neuroblast-related. Analysis of the gene-set linked to concordant loci identified the biological process, “presynaptic active zone organization” and three cell type signatures, HGABA and HNBGABA in the midbrain as well as “fetal limbic system neurons” (Table S7). The gene-set linked to discordant loci identified many more mechanisms including 11 biological processes and 14 cell-type signatures, though the latter did not include limbic system neurons (Table S8).

### Drug repurposing analysis

Of the 131 unique genes associated with the 132 lead SNPs, six (*OPRM1*, *RET*, *DPYD*, *ADH1A*, *PDE4B*, *PRKCB*) were located in the gene or nearest to the transcription start site of genes associated with FDA-approved drugs (i.e., Tclin; Table S4), including naltrexone (*OPRM1*) among others. Eleven other genes were Tchem (i.e., known to bind to small molecules with high potency), followed by 101 Tbio and 13 Tdark^44^. Two of the 11 Tchem targets are targeted by known drugs in OpenTargets. Specifically, *FTO* is targeted by bisantrene for acute myeloid leukemia (Phase II), and *GRM2* is targeted by four investigational drugs for central nervous system disorders including schizophrenia, major depressive disorder, perceptual disorders, bipolar disorder, psychosis, methamphetamine dependence, post-traumatic stress disorder, and seizure disorder.

### BrainXcan

AUD was significantly associated with gray matter volume in four subcortical brain regions after applying p-value corrections for LD structure and multiple testing: the bilateral caudate nucleus (positive association), left amygdala (negative association), and right thalamus (positive association) (Table S9, Figures S6 and S7). After p-value adjustments, BMI had a total of 98 significant associations with brain IDPs. Approximately three-quarters of the BMI associations were with diffusion MRI IDPs (n = 74; 75.51%), with the remainder coming from structural MRIs. The top associations for BMI were with the medial lemniscus, a white matter tract that is part of the somatosensory pathway responsible for transmitting tactile, proprioceptive, and other sensations from the body to higher brain centers^56^. All significant associations for AUD had the same direction of effect for BMI (Figure 5), 76 of the BMI associations had the same direction of effect as AUD, and 22 had effects in the opposite direction for the two traits (Table S9, Figures S8 and S9). Gray matter volume in three brain regions—the bilateral caudate and the left amygdala—was significantly associated with both AUD and BMI.

**Figure 5.**
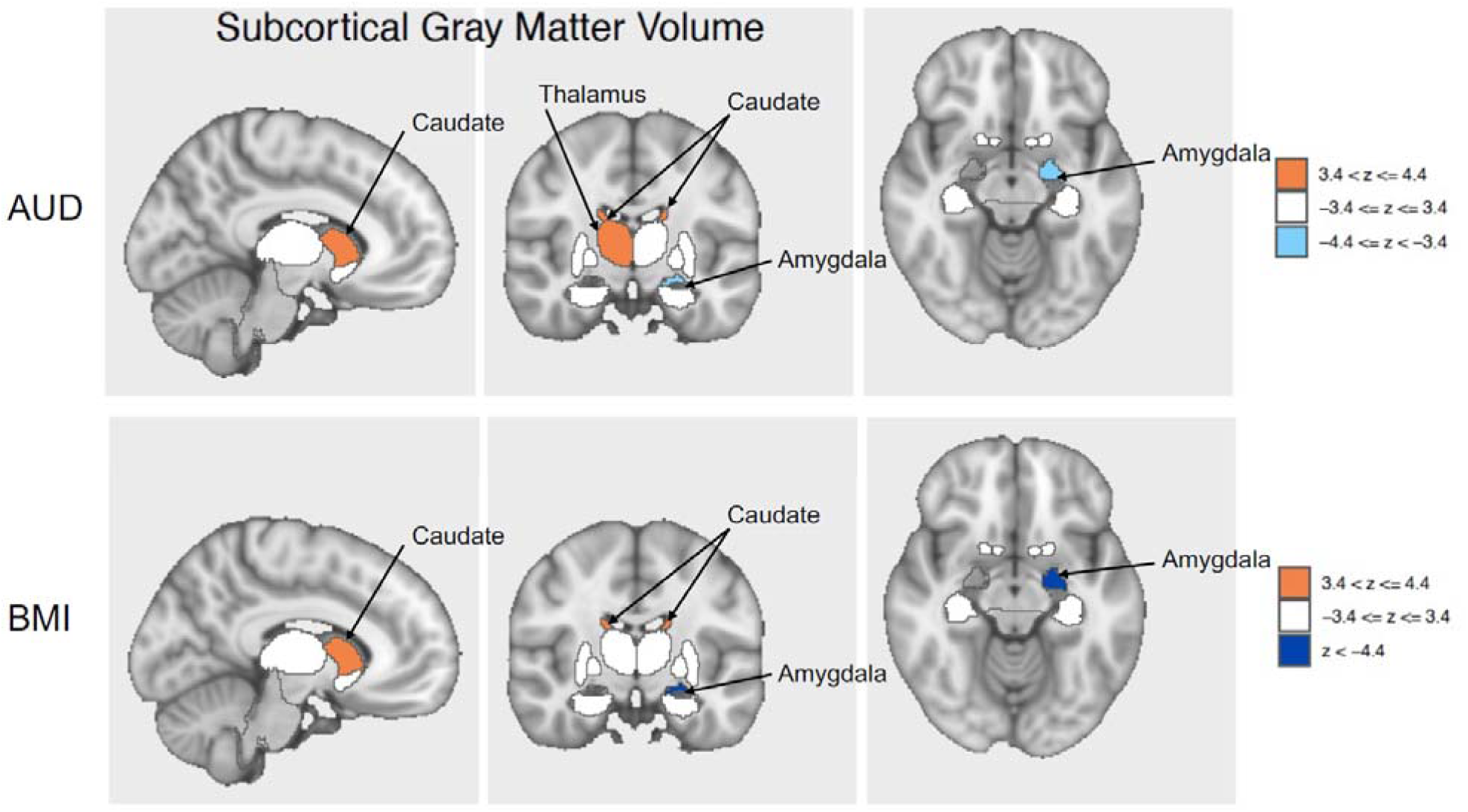
Brain visualization of subcortical features significantly associated with AUD and BMI. Z-scores of the associations of the brain regions significant after false discovery rate correction (bilateral caudate, right thalamus, left amygdala) are shown, with orange indicating positive associations and blue indicating negative associations. AUD = alcohol use disorder, BMI = body mass index.

## Discussion

Analyses revealed several key findings regarding the genetic relationship between AUD and BMI. Both MiXeR and conjFDR analyses showed that more than half the variants shared by AUD and BMI exerted opposite directions of effect on the traits, supporting our hypothesis that this underlies their lack of genetic correlation. Leveraging conjFDR, we identified 132 shared genomic loci, including 53 that were novel from the original GWAS for AUD and BMI. Furthermore, analyses in a smaller independent sample demonstrated high replicability of the sign concordance of these results and moderate replicability of the significant associations, enhancing confidence in the findings. Expression analysis of genes linked to both phenotypes identified heightened expression in brain regions implicated in executive functioning, reward, homeostasis, and food intake regulation. BrainXcan analyses of brain IDPs from the UK Biobank reinforced these findings, identifying significant shared associations with caudate nucleus and amygdala. Overall, these findings detail the extensive polygenic overlap between AUD and BMI, elucidate several overlapping neurophysiological mechanisms, and suggest possible targets for intervention.

The low genetic correlation found in the present and prior studies^22,23^ is explained by the mixed directionality of genetic effects of the phenotypes. MiXeR analysis indicated high polygenic overlap (82%) between AUD and BMI, with 49% concordance in the variants’ effect directions. Similarly, using conjFDR, 42.4% (n = 56) of the 132 loci significantly associated with both AUD and BMI had the same direction of effect. Comparison analyses examining the overlap of AUD and BMI with other psychiatric phenotypes (ADHD, MDD, schizophrenia) yielded a range of genetic correlations and effect direction concordance, all showing substantial genetic overlap, consistent with prior work^23,57,58^. AUD and BMI exhibited the highest proportion of estimated shared causal variants of all other phenotype pairings, despite having the lowest absolute genetic correlation of any pairing, which underscores the importance of accounting for variants’ effect direction. The LAVA analysis corroborated these findings, revealing that 41 of the 2,495 independent regions exhibited significant genetic correlations between AUD and BMI, with nearly half (19 out of 41) showing positive correlations.

The *FTO* and *SLC39A8* gene loci exhibited the most significant discordant effects, and the *CADM2* gene locus exhibited the most significant concordant effects. Specifically, the effect allele of rs9939973, intronic within the *FTO* gene, was associated with a reduced likelihood of AUD and increased BMI. While BMI and alcohol-related GWAS have identified intronic FTO variants^23,59,60^, this conjunctional analysis highlights this locus as the region that is most significantly associated with both phenotypes. Obesity research suggests that the *FTO* gene region alters the function of nearby genes (*IRX3* and *IRX5*), which impact the involvement of fat cells in thermogenesis ^61,62^. The psychiatric literature suggests that this region also affects neuronal activity, namely dopamine receptor type 2 and 3 function ^63,64^. The *SLC39A8* gene, which encodes the metal ion transporter protein ZIP8 ^65^, was associated with both AUD and BMI, but with opposite directions of effect. This locus is highly pleiotropic and has been linked to an array of psychiatric and neurological diseases and Crohn’s disease, possibly due its role in maintaining manganese homeostasis^47,66,67^. Conversely, the most significant variant with concordant effects on AUD and BMI was rs10511087, intronic within the *CADM2* gene. *CADM2* is expressed in some brain regions implicated in the present study, namely the frontal cortex (BA9) (Figure 3), and has been associated with cognition, pain, impulsivity, substance use, other risky behaviors, obesity, and other metabolic traits ^48,49,68–70^.

Drug repurposing analysis yielded targets of FDA-approved drugs and others that are in development or have not been examined. In particular, *PDE4B*, which is targeted by FDA-approved medications for chronic obstructive pulmonary disease and psoriasis/psoriatic arthritis, has shown evidence of reducing alcohol consumption^71,72^ and is under investigation as a treatment target for AUD (NCT05414240), weight loss, and other metabolic conditions^73–75^. Additionally, *OPRM1* is targeted by naltrexone, which is FDA-approved for treating AUD and opioid use disorder, and as a combination drug, naltrexone-bupropion, for the treatment of obesity^19,76^ Importantly, both the *PDE4B* and *OPRM1* loci exhibited concordant effects on AUD and BMI. However, studies have not examined the effectiveness of these medications in treating both conditions simultaneously in a clinical setting and this study underscores the relevance of examining treatment options focused on genes linked to both phenotypes.

A final promising target identified in this study is, *GRM2,* which encodes for the metabotropic glutamate receptor 2 (mGluR2) and exhibited a discordant effect between AUD and BMI in the present study. Both metabotropic glutamate receptor (mGluR) 2/3 agonists and mGluR2 positive allosteric modulators have been found to reduce drinking after alcohol deprivation^77^, alcohol self-administration^78,79^, cue-induced reinstatement^78^, and stress-induced reinstatement^79^ (for a review of preclinical alcohol studies see ^80^). Administration of an mGluR2 agonist has also been shown to simultaneously reduce ethanol- and sucrose-seeking, and body weight^81^. It will be important for future studies to investigate the discordant effect identified in the present study and further evaluate the promise of this mechanism for the treatment of AUD and obesity.

Several brain regions and neural cell types were identified across the various downstream analyses. Genes linked to the lead SNPs were significantly up-regulated in the prefrontal and anterior cingulate cortex, hypothalamus, hippocampus, and amygdala. Additionally, amygdala and caudate nucleus gray matter volumes were significantly associated with AUD and BMI risk in BrainXcan analyses. Furthermore, GO gene-set analysis yielded enrichment for signatures in the midbrain, including GABAeric, dopaminergic, and serotonergic cell types. Shared genes linked to concordant loci identified up-regulation in the amygdala and hypothalamus and implicated limbic system neurons, whereas discordant loci genes implicated the prefrontal and anterior cingulate cortex. The brain regions and cell types identified here have consistently been implicated in obesity^82,83^, binge eating disorder^84,85^, and AUD^86,87^. The prefrontal cortex and amygdala are dysregulated after drug use, resulting in withdrawal, craving, impulsivity, and negative affect, which drive continued use^87,88^. Similarly, the frontal cortex, amygdala, caudate, and hypothalamus exhibit hyperreactivity to food-associated cue exposure in obese and overweight individuals^83^. Evidence also suggests that the caudate nucleus plays a key role in mediating external stimuli and internal preferences to guide behavior^89^. The hypothalamus is integral to stress responses and homeostatic regulation of caloric intake to meet real and perceived nutrition needs^90,91^. Dysfunction in these areas is progressive with eating and drug use, resulting in altered reward processing and a shift in ‘liking’ vs. ‘wanting’ the hyper-fixated substance^92–94^. Overall, these findings underscore the utility of leveraging human genomic and transcriptomic data analysis and align with insights from preclinical and human neuroimaging studies on the neurophysiological mechanisms driving AUD and obesity. Future studies leveraging high resolution single-cell RNA sequencing datasets^95^ are needed to parse the brain regions implicated and understand the mechanisms underlying concordant and discordant shared loci.

There are notable limitations to this study. BMI may not optimally measure obesity, both because it is used as a continuous variable in this study, and because there is no distinction between weight from fat, bone, or muscle mass. However, BMI is easily and inexpensively measured, and is a longstanding, well-studied surrogate measure of obesity ^96^. Nonetheless, future studies should expand these analyses to examine complementary phenotypes, such as body composition, waist-to-hip ratio^97^, and binge-eating disorder^98^, and to clinical subpopulations (e.g., through GWAS of BMI in individuals with and without AUD) ^99^.

A second limitation is that the data used in this study are limited to individuals of European ancestry to ensure compatible genetic architectures. As sample sizes increase, a high priority must be placed on including individuals of non-European ancestry. The potential consequences of excluding diverse ancestral groups include the inequitable distribution of the benefits of genetic research and exacerbation of existing health disparities^100–102^. Thus, future studies should replicate these findings using more diverse ancestral samples as biobanks continue to grow. This shift towards more diverse samples will improve result generalizability and understanding of cross-population genetic variation.

Finally, while the use of data aggregated from large biobank studies enables highly powered genomic analyses, there is limited ability to account for environmental exposures and longitudinal progression. Family studies such as the Collaborative Study on the Genetics of Alcoholism (COGA)^103^ can provide complementary data to larger-scale biobank studies. Indeed, follow-up studies leveraging data from studies like COGA will enable a better understanding of AUD and BMI progression, remission and recovery, antecedents and sequelae, and the interplay between genetic and socio-environmental factors.

In summary, our study found that the absence of genetic correlation between AUD and BMI is attributable to the presence of shared variants with opposite directions of effect (i.e., a variant protective for obesity increases risk for AUD and vice versa). Follow-up analyses specified overlapping genomic loci and identified brain regions implicated in executive functioning, reward, homeostasis, and food intake regulation. Together, these findings increase our understanding of the shared polygenic architecture of AUD and BMI and lend further support to the notion that eating behavior and AUD share overlapping neurobiological mechanisms.

## Data availability

Full summary statistics from two genome-wide association studies can be accessed at the following locations: GIANT Consortium website (https://portals.broadinstitute.org/collaboration/giant/index.php/GIANT_consortium_data_files) and through the Gelernter Lab website without restriction (https://medicine.yale.edu/lab/gelernter/stats/) or dbGaP (accession number phs001672, under the ‘Addiction’ Analysis; registration and approval are needed following dbGaP’s data accessing process). Researchers seeking access to the exact AUD summary statistics cohort used in this study should contact the original study authors for more information (Zhou et al., 2023). Replication summary statistics can be accessed at the FinnGen website (https://www.finngen.fi/en/access_results).

## Code availability

MiXeR (https://github.com/precimed/mixer) was used to investigate the overall shared genetic architecture between AUD and BMI. LD Score regression (LDSC) v1.0.1 (https://github.com/bulik/ldsc) was used to calculate heritability, genetic correlations, and standard errors. conjFDR (https://github.com/precimed/pleiofdr) analysis was used to detect loci significantly associated with both phenotypes. Local Analysis of [co]Variant Association (LAVA) was used to estimate local heritability and local genetic correlations (https://github.com/josefin-werme/LAVA). Functional Mapping and Annotation (FUMA) was used to identify lead single nucleotide polymorphisms and to implement gene expression, tissue enrichment specificity, and gene-set enrichment analyses. FUMA can be accessed here: https://fuma.ctglab.nl. Annotations for the lead SNPs corresponding to Variant Effect Predictor (VEP), Combined Annotation Dependent Deletion (CADD) scores, and nearest transcription start site were sourced from OpenTargets (https://genetics.opentargets.org/ v22.10). The presence of lead SNPs within genes was confirmed using dbGaP (https://www.ncbi.nlm.nih.gov/gap/). The Target Central Resource Database (TCRD) and OpenTargets were accessed to integrate drug-protein interaction/druggability information (https://pharos.nih.gov/ v3.18.0; https://platform.opentargets.org/v23.12). BrainXcan was used to evaluate the shared associations of AUD and BMI with brain image-derived phenotypes. BrainXcan documentation can be found here: https://liangyy.github.io/brainxcan-docs/docs/overview.html.

## Funding Source

This work was supported by funding from the National Institute on Alcohol Abuse and Alcoholism (R01 AA030041 and R01 AA030056), the Department of Defense (HU0001-22-2-0066), and the Veterans Integrated Service Network 4 Mental Illness Research, Education and Clinical Center of the Crescenz Veterans Affairs Medical Center. LL is a federal employee and is supported by the National Institute on Drug Abuse and the National Institute on Alcohol Abuse and Alcoholism Intramural Research Programs.

## Disclaimer

The opinions and assertions herein are those of the authors and do not necessarily reflect the official views of the Henry M. Jackson Foundation for the Advancement of Military Medicine, Inc. Moreover, the opinions and assertions herein do not necessarily reflect the official views of the Department of Defense, Uniformed Services University, the National Institute on Alcohol Abuse and Alcoholism, the US Government, and do not imply endorsement by the Federal Government.

## Disclosures

HRK is a member of advisory boards for Altimmune, Clearmind Medicine, Dicerna Pharmaceuticals, Enthion Pharmaceuticals, Lilly Pharmaceuticals, and Sophrosyne Pharmaceuticals; a consultant to Sobrera Pharmaceuticals and Altimmune; the recipient of research funding and medication supplies for an investigator-initiated study from Alkermes; and a member of the American Society of Clinical Psychopharmacology’s Alcohol Clinical Trials Initiative, which was supported in the last three years by Alkermes, Dicerna, Ethypharm, Imbrium, Indivior, Kinnov, Lilly, Otsuka, and Pear. JG and HRK hold U.S. patent 10,900,082 titled: “Genotype-guided dosing of opioid agonists,” issued 26 January 2021. JG is paid for editorial work for the journal *Complex Psychiatry*.

---

**Figure S1.**
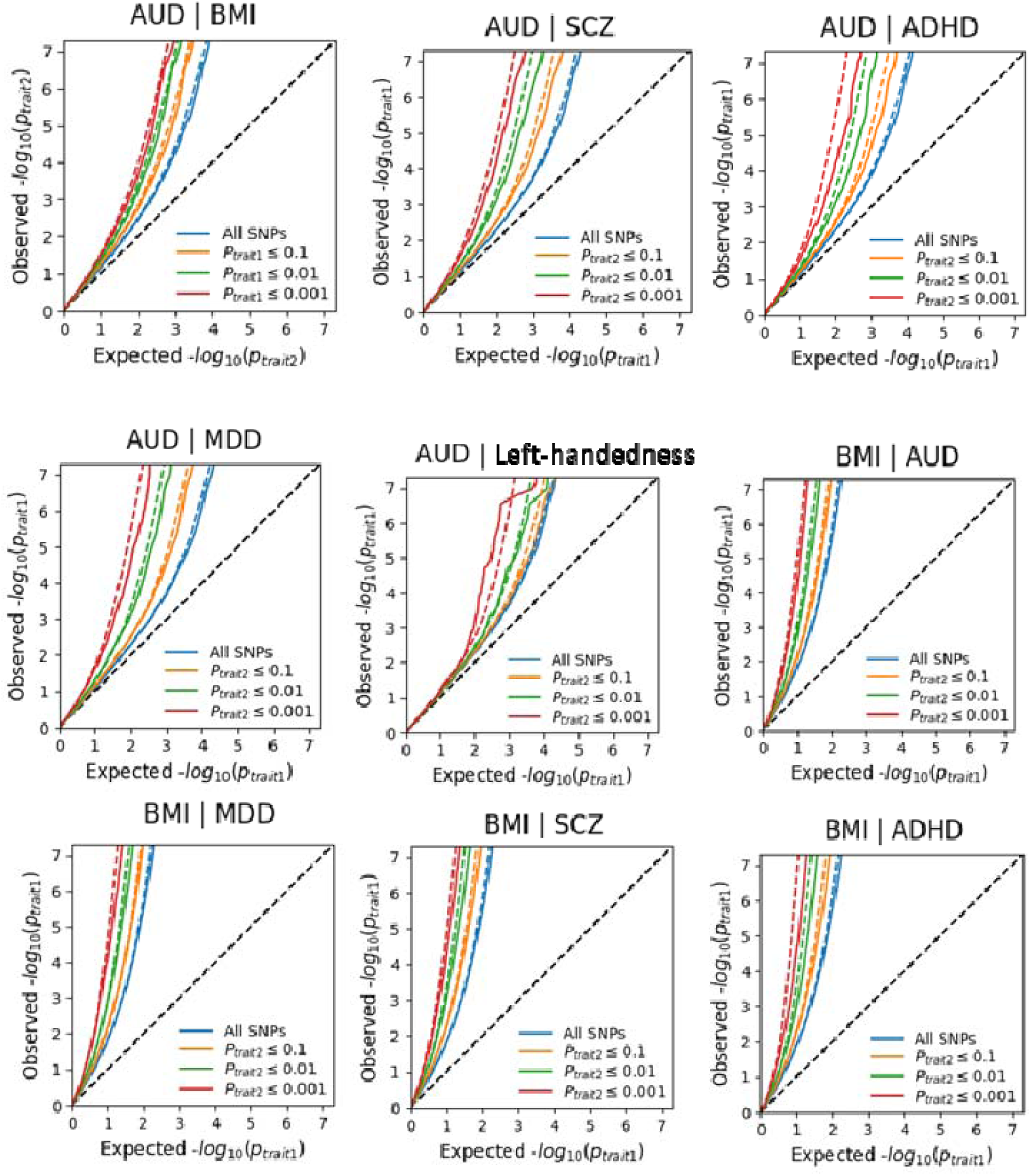
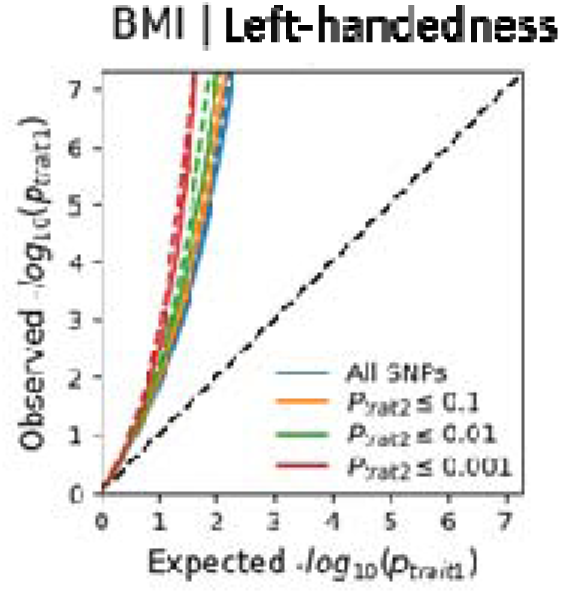
Conditional Q-Q plots showing the distribution of observed versus expected –log_10_ *p*-values for the primary phenotypes (AUD and BMI) for SNPs conditional on associations of a secondary trait at three p-value strata (*p* ≤ 0.1, 0.01, and 0.001). ADHD = attention deficit hyperactivity disorder, AUD = alcohol use disorder, BMI = body mass index, MDD = major depressive disorder, SCZ = schizophrenia.

**Figure S2.**
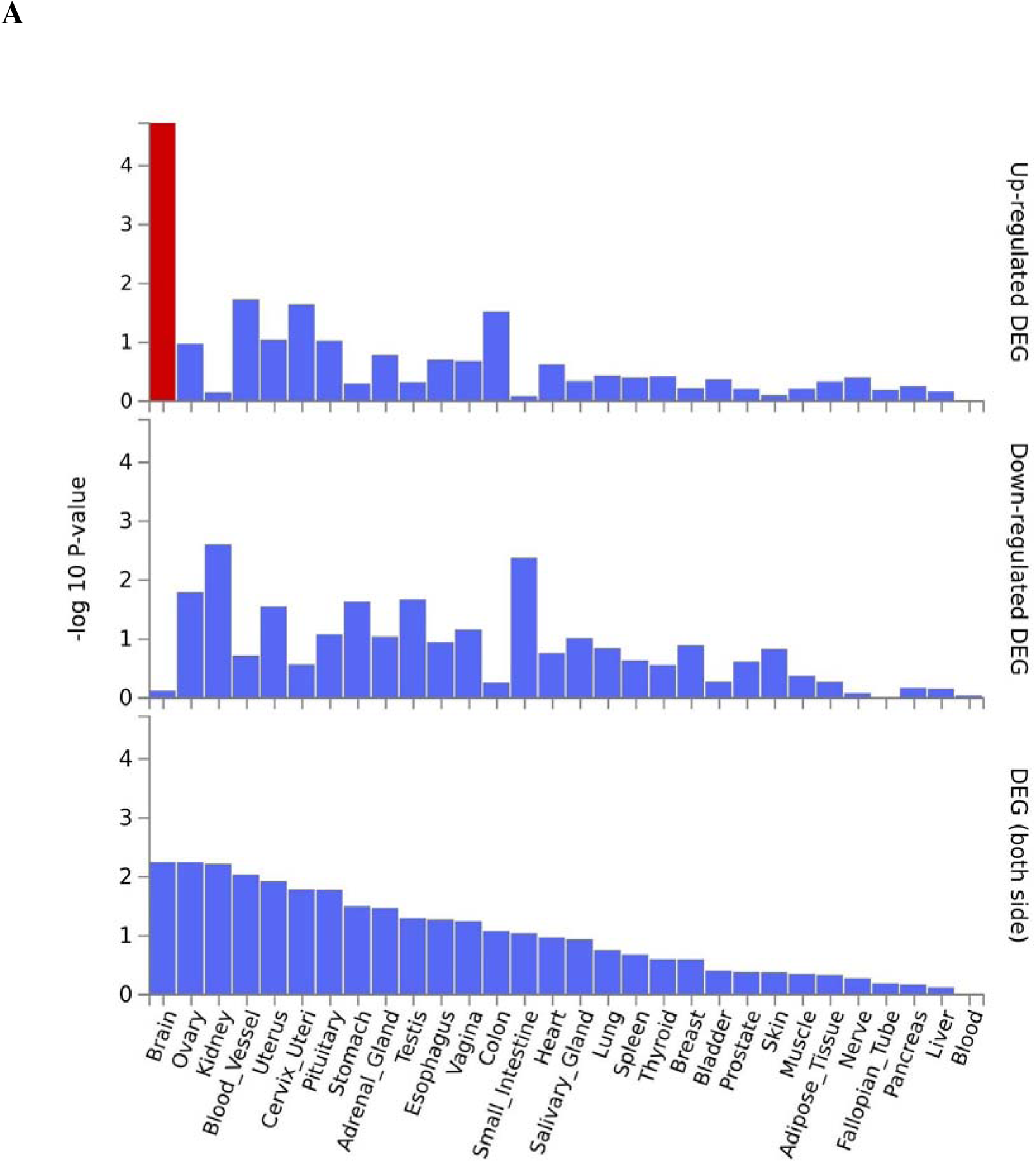
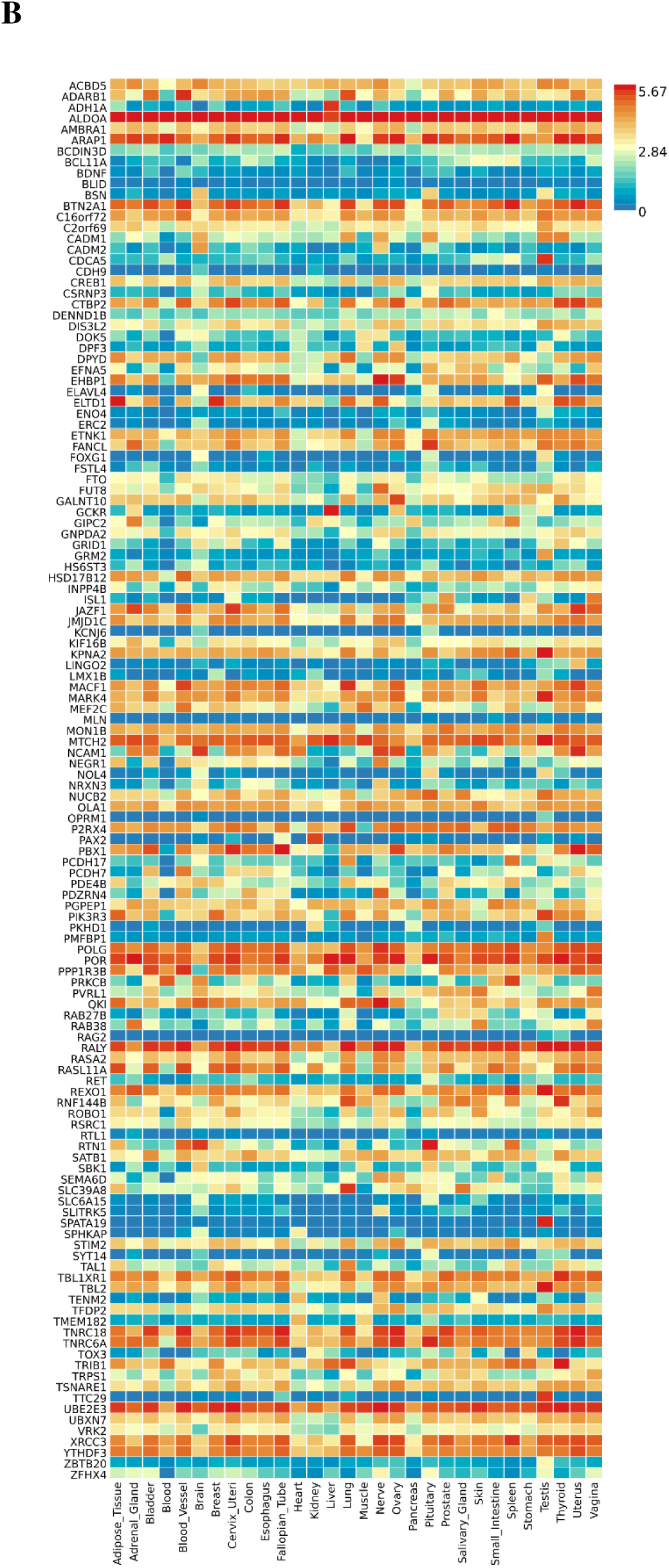
**A)** Differential gene expression (DEG) in 30 general GTEx tissues for genes linked to lead SNPs in distinct loci significantly associated with both AUD and BMI. Significant enrichment (p<.05 after Bonferroni correction) is highlighted in red. **B)** Gene expression heatmap for 30 GTEx general tissues for genes linked to lead SNPs in distinct loci significantly associated with both AUD and BMI.

**Figure S3.**
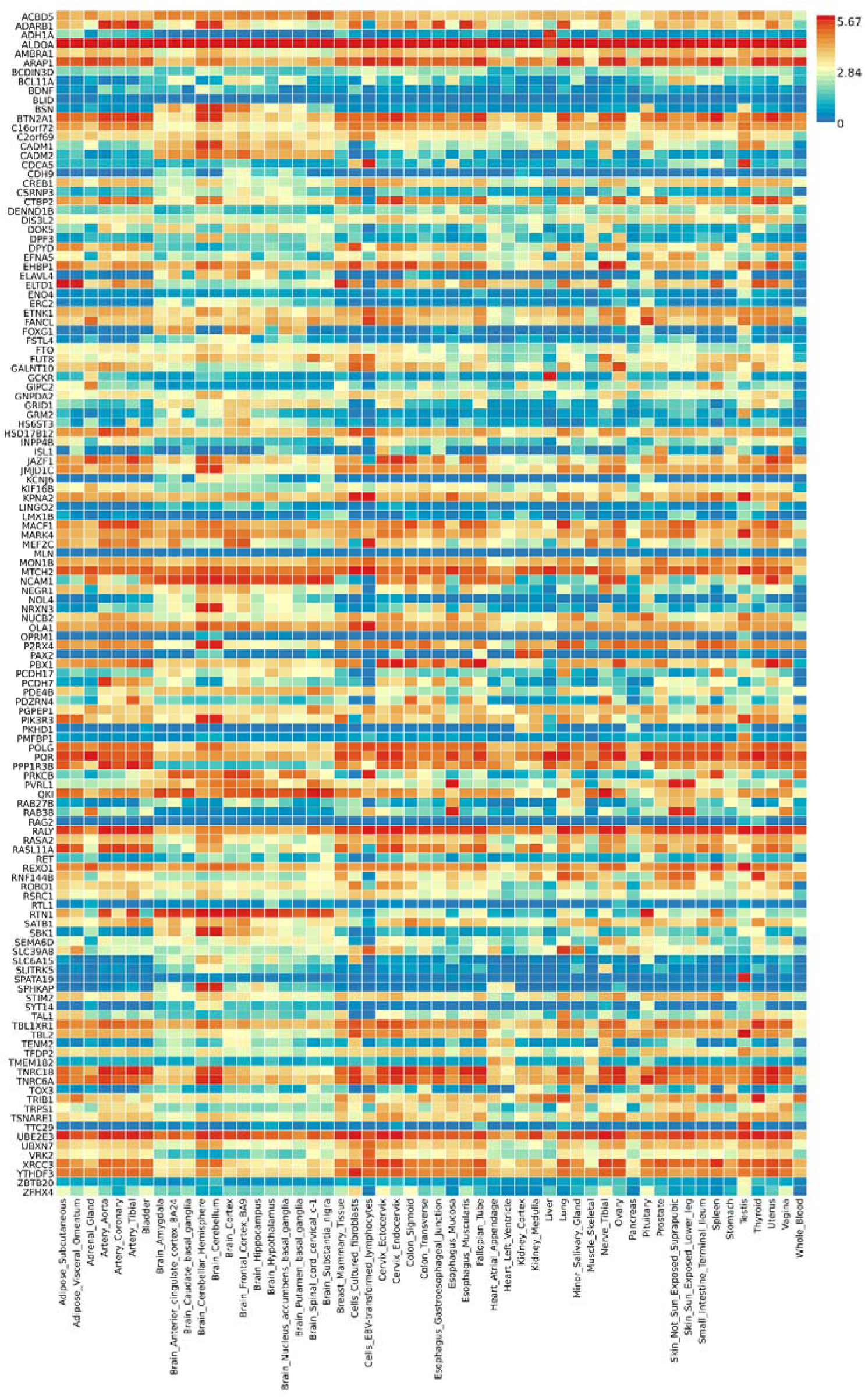
Gene expression heatmap for 54 GTEx tissues for genes linked to lead SNPs in distinct loci significantly associated with both AUD and BMI.

**Figure S4.**
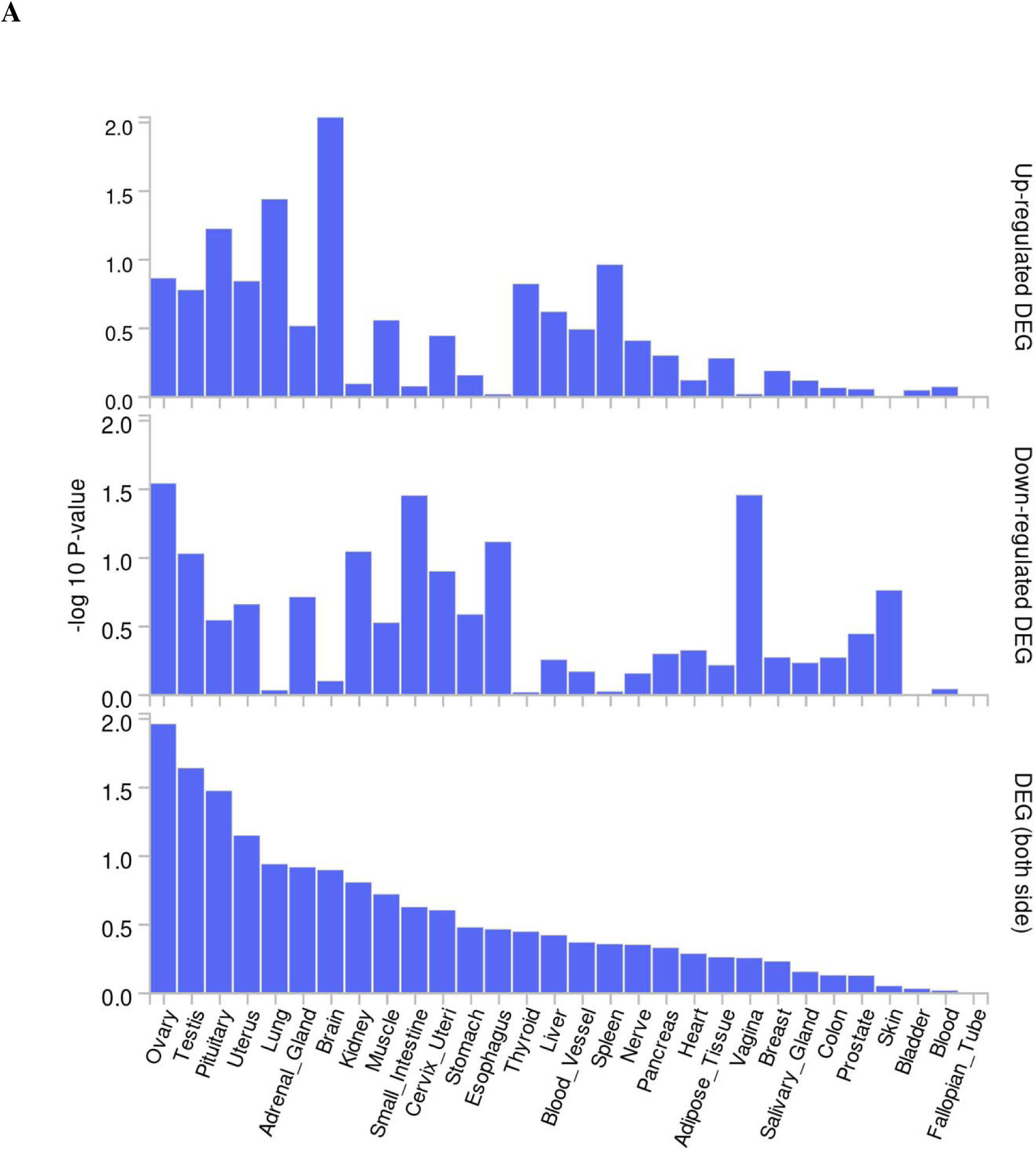
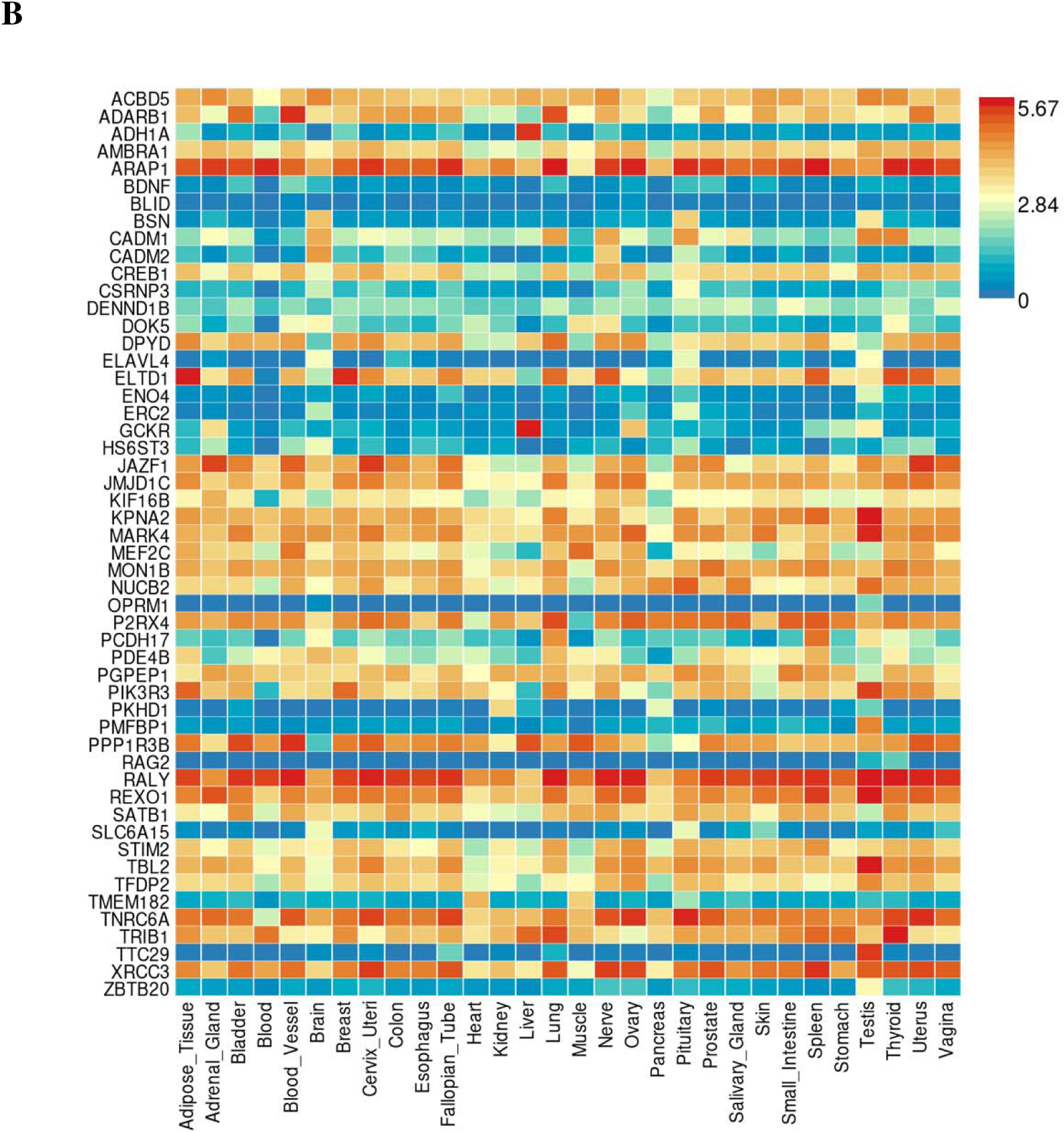
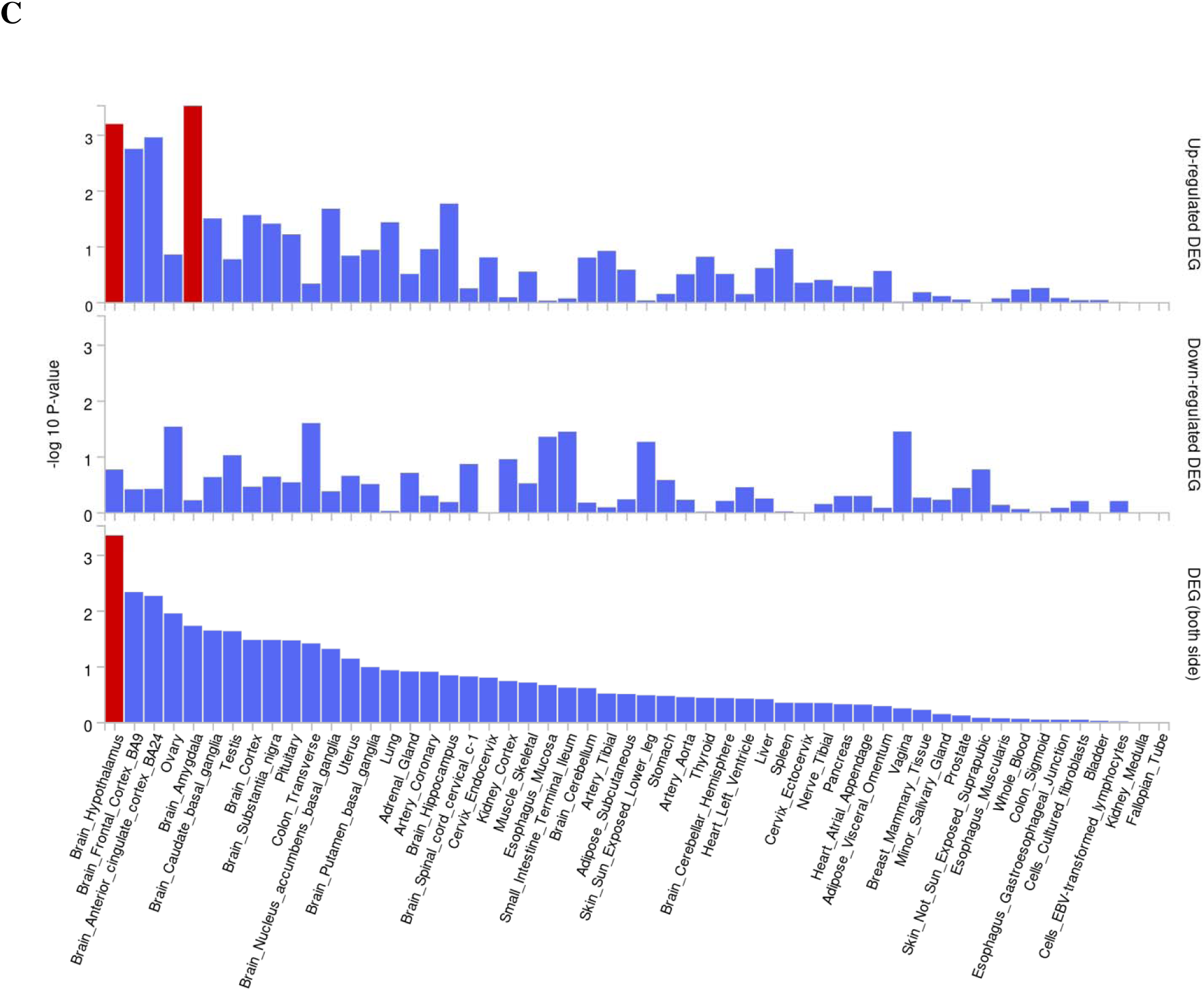
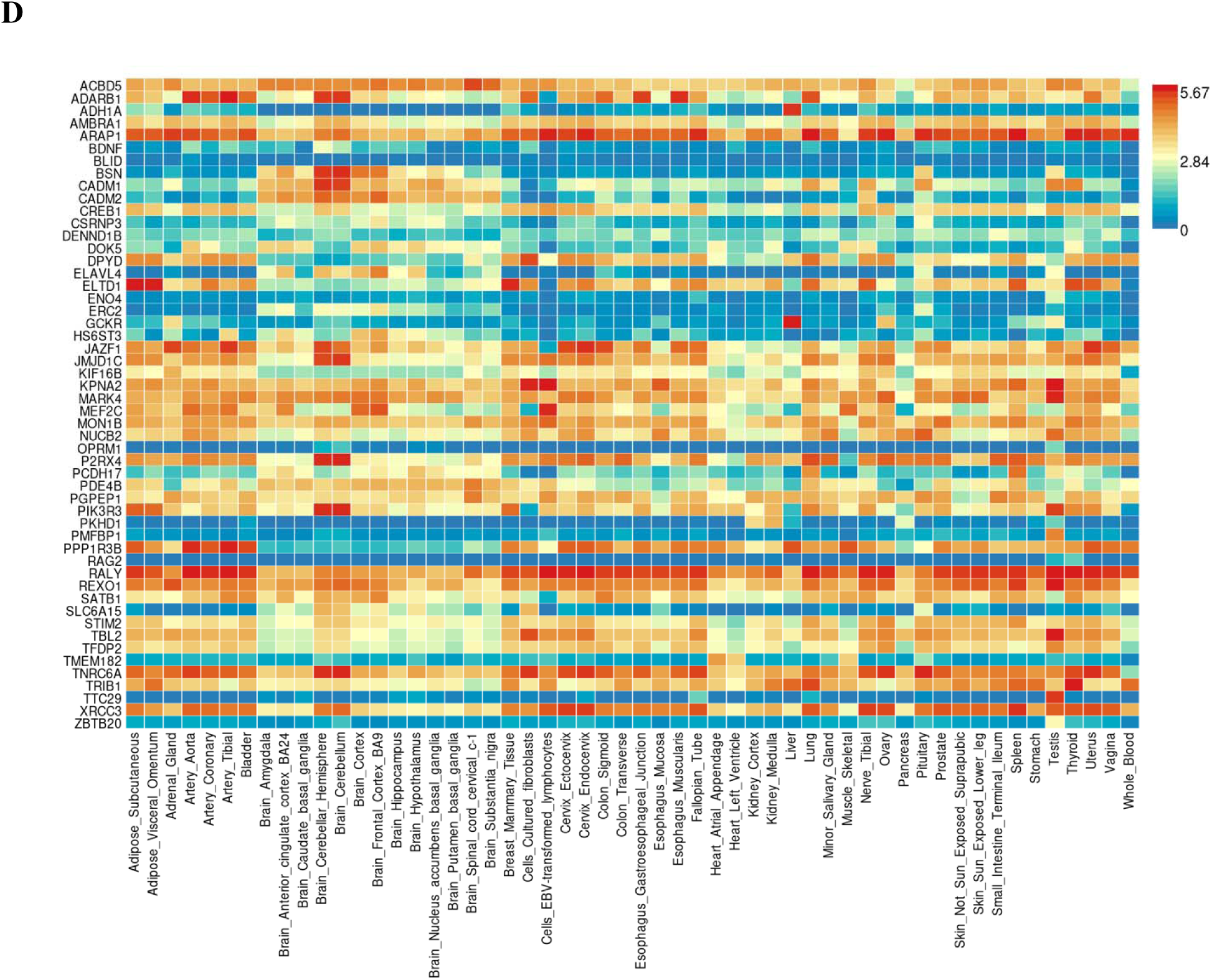
Concordant enrichment. **A)** Differential gene expression (DEG) in 30 general GTEx tissues for genes linked to concordant lead SNPs in distinct loci significantly associated with both AUD and BMI. There was no significant enrichment (p<.05 after Bonferroni correction). **B)** Gene expression heatmap for 30 GTEx general tissues for genes linked to concordant lead SNPs in distinct loci significantly associated with both AUD and BMI. **C)** Differential gene expression (DEG) in 54 general GTEx tissues for genes linked to concordant lead SNPs in distinct loci significantly associated with both AUD and BMI. Significant enrichment (p<.05 after Bonferroni correction) is highlighted in red. **D)** Gene expression heatmap for 54 GTEx general tissues for genes linked to concordant lead SNPs in distinct loci significantly associated with both AUD and BMI.

**Figure S5.**
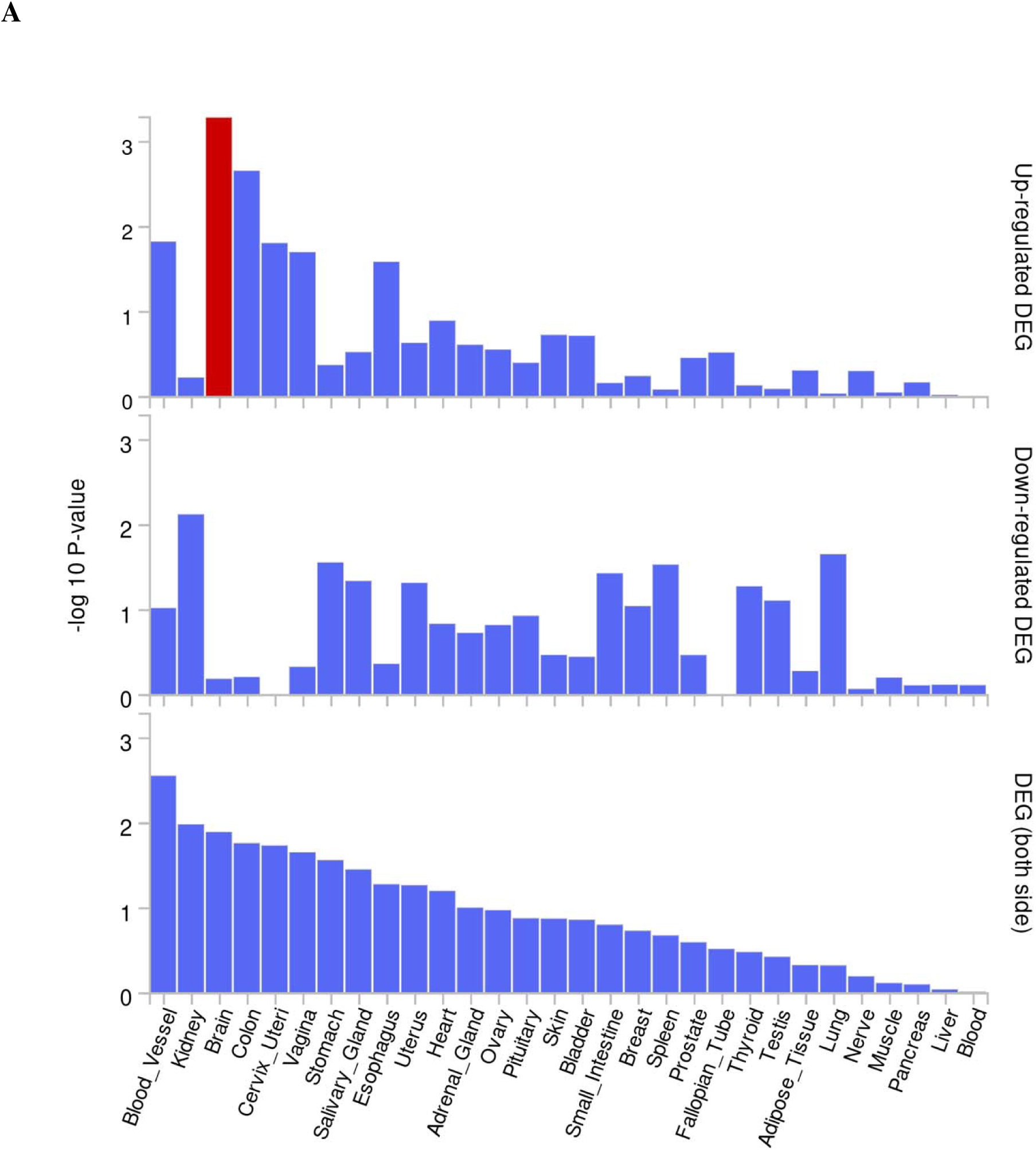
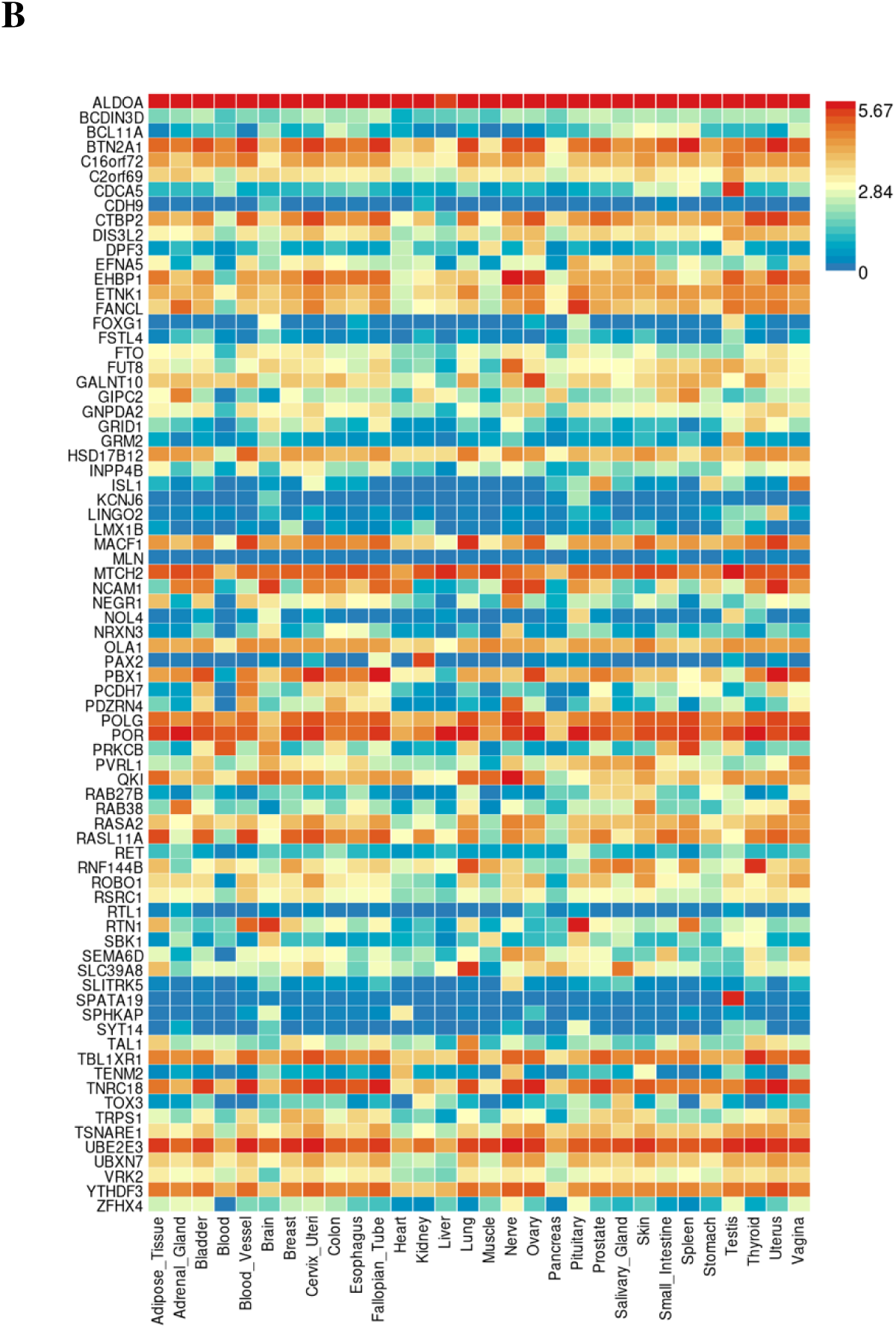
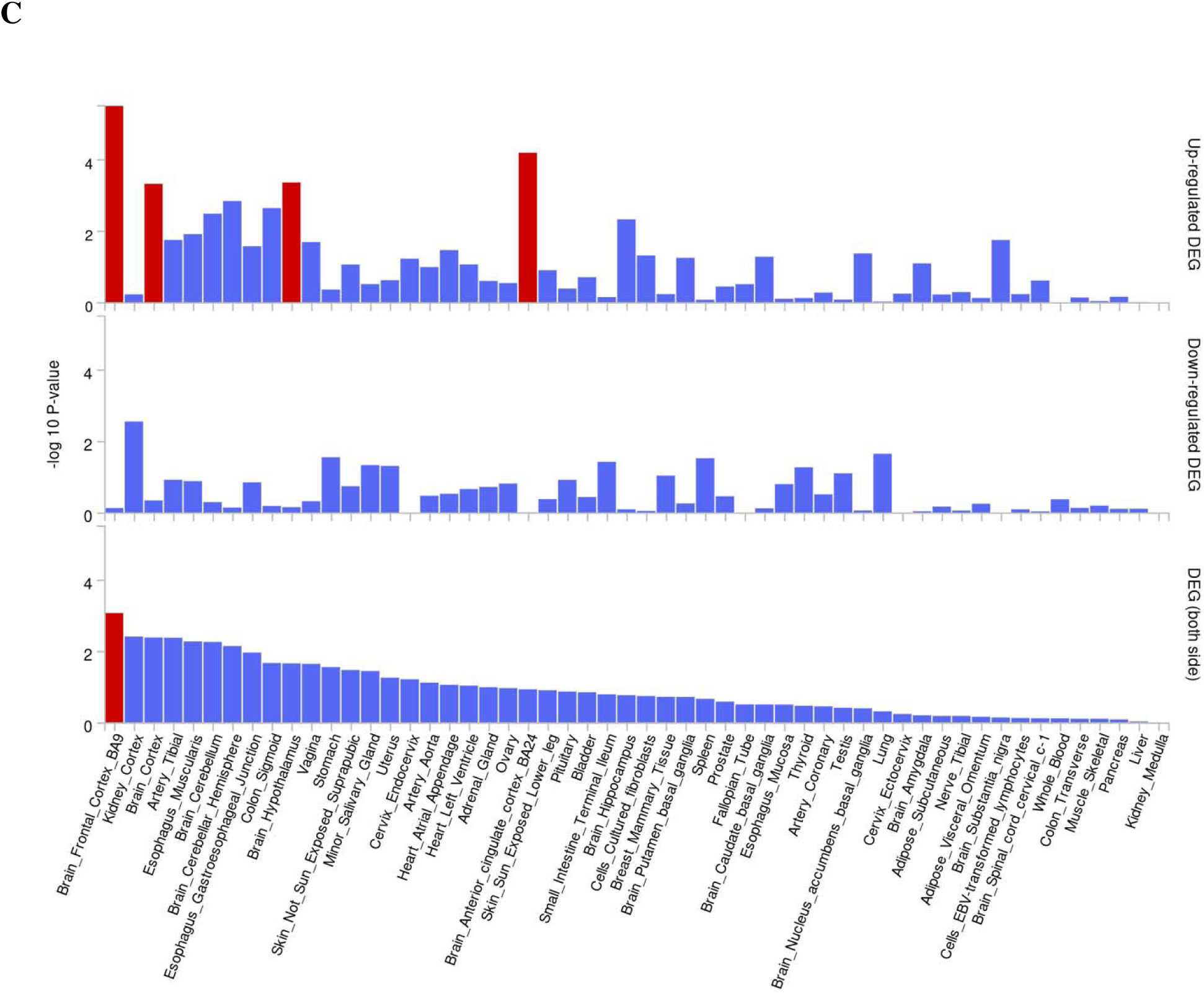
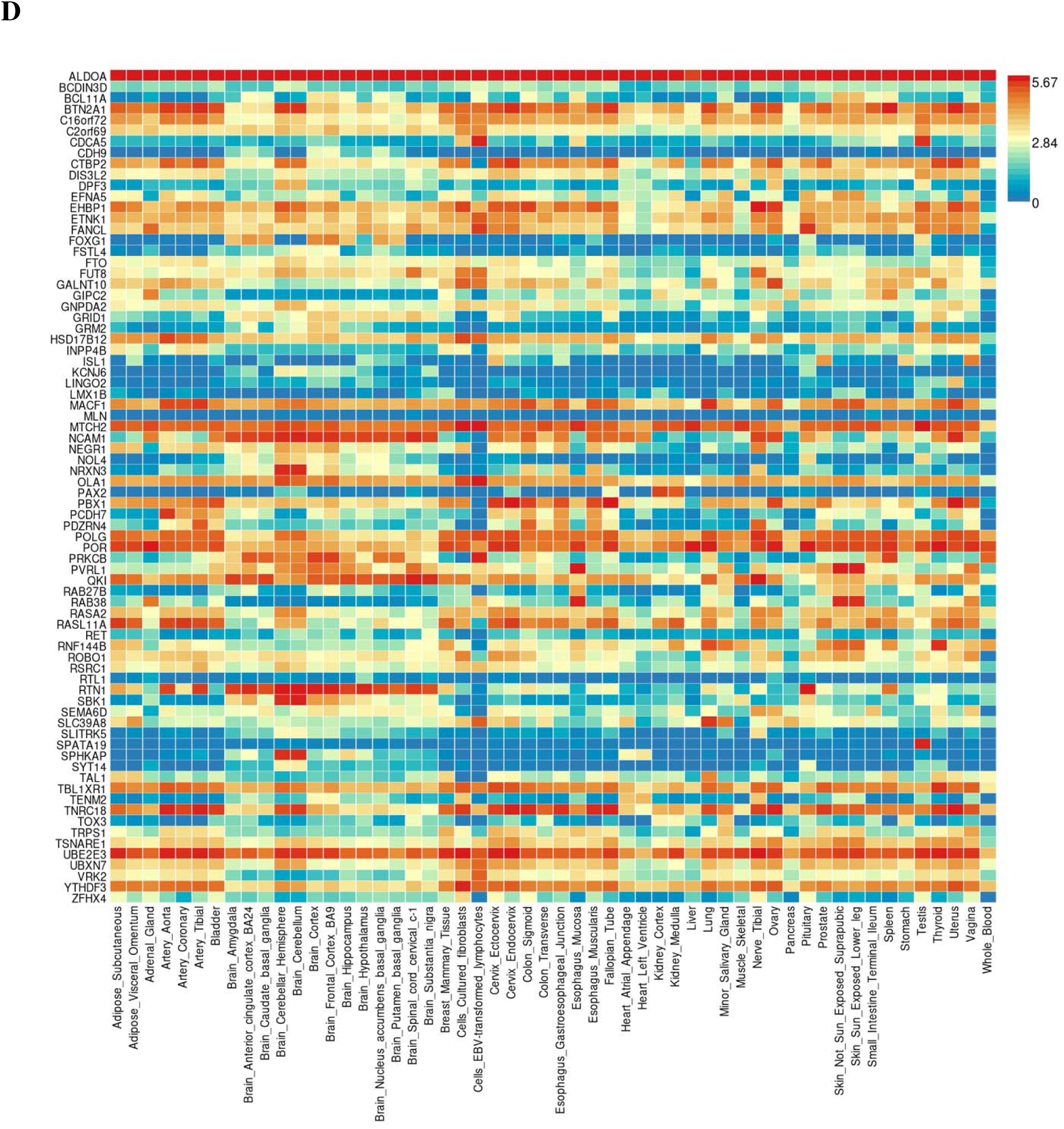
Discordant enrichment. **A)** Differential gene expression (DEG) in 30 general GTEx tissues for genes linked to discordant lead SNPs in distinct loci significantly associated with both AUD and BMI. There was no significant enrichment (p<.05 after Bonferroni correction). **B)** Gene expression heatmap for 30 GTEx general tissues for genes linked to discordant lead SNPs in distinct loci significantly associated with both AUD and BMI. **C)** Differential gene expression (DEG) in 54 general GTEx tissues for genes linked to discordant lead SNPs in distinct loci significantly associated with both AUD and BMI. Significant enrichment (p<.05 after Bonferroni correction) is highlighted in red. **D)** Gene expression heatmap for 54 GTEx general tissues for genes linked to discordant lead SNPs in distinct loci significantly associated with both AUD and BMI.

**Figure S6.**
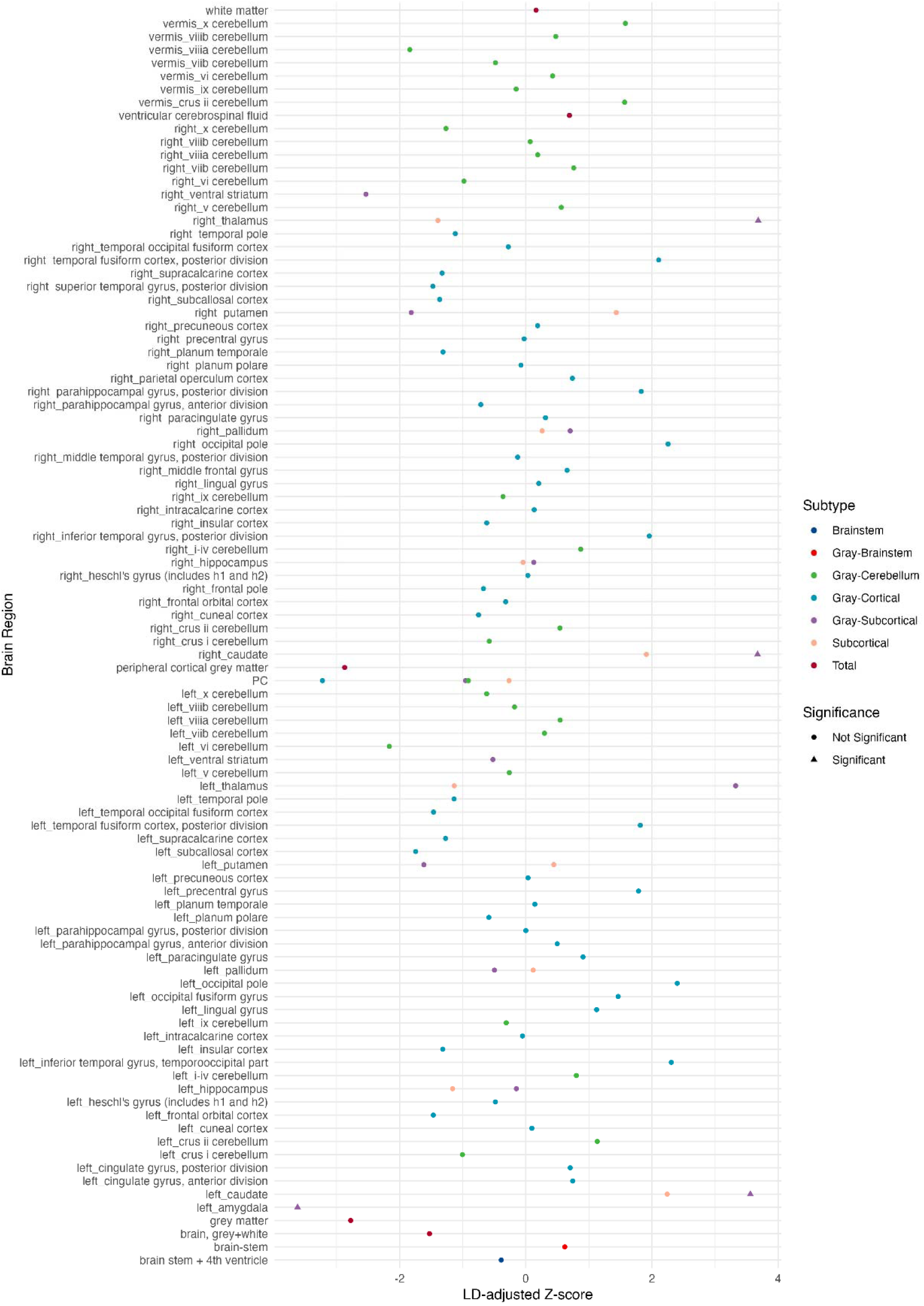
Structural brain features associated with AUD risk. LD-adjusted Z-scores are provided for region-specific associations of structural IDPs with AUD risk.

**Figure S7.**
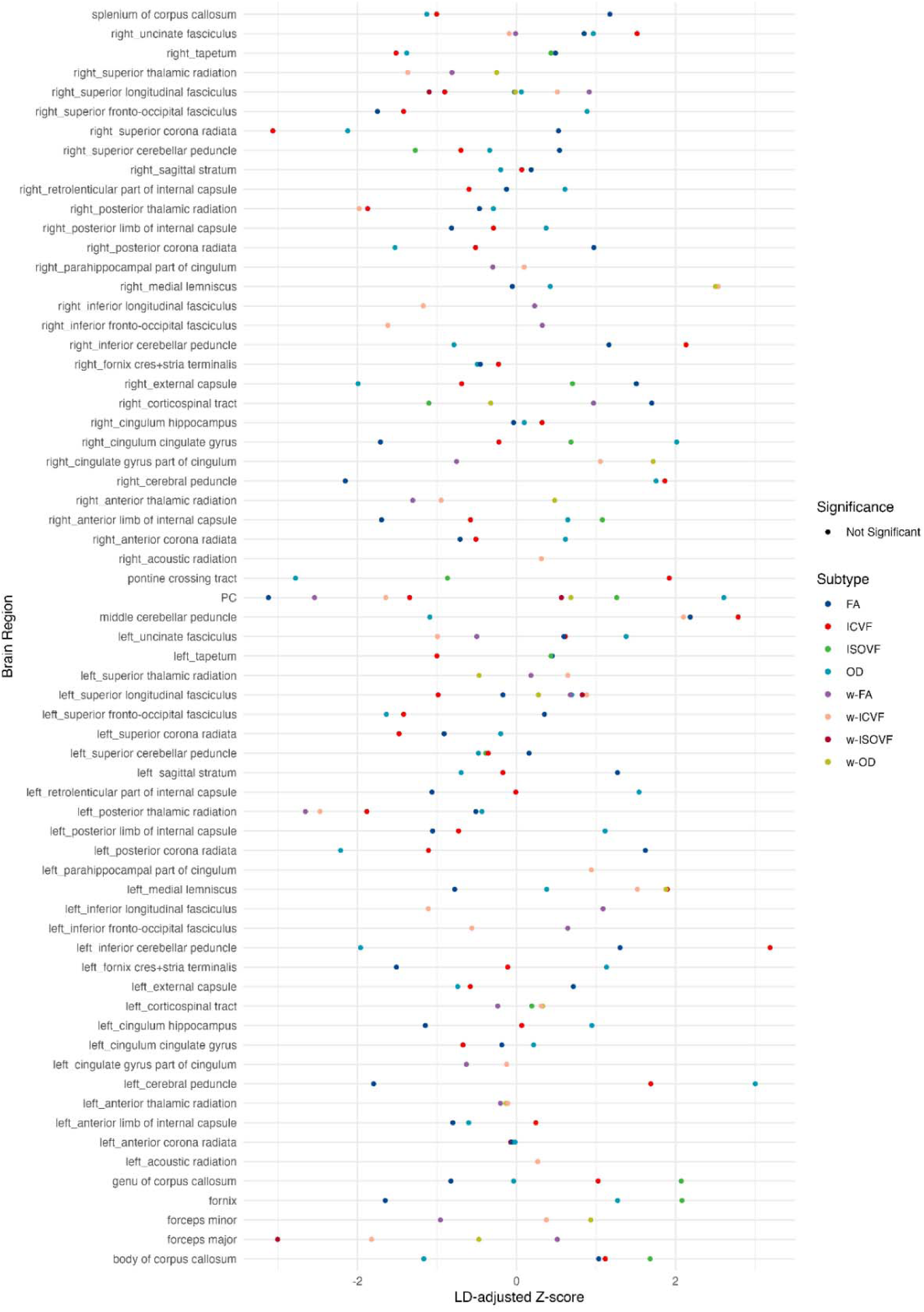
Diffusion features associated with AUD risk. LD-adjusted Z-scores are provided for region-specific associations of diffusion MRI IDPs with AUD risk.

**Figure S8.**
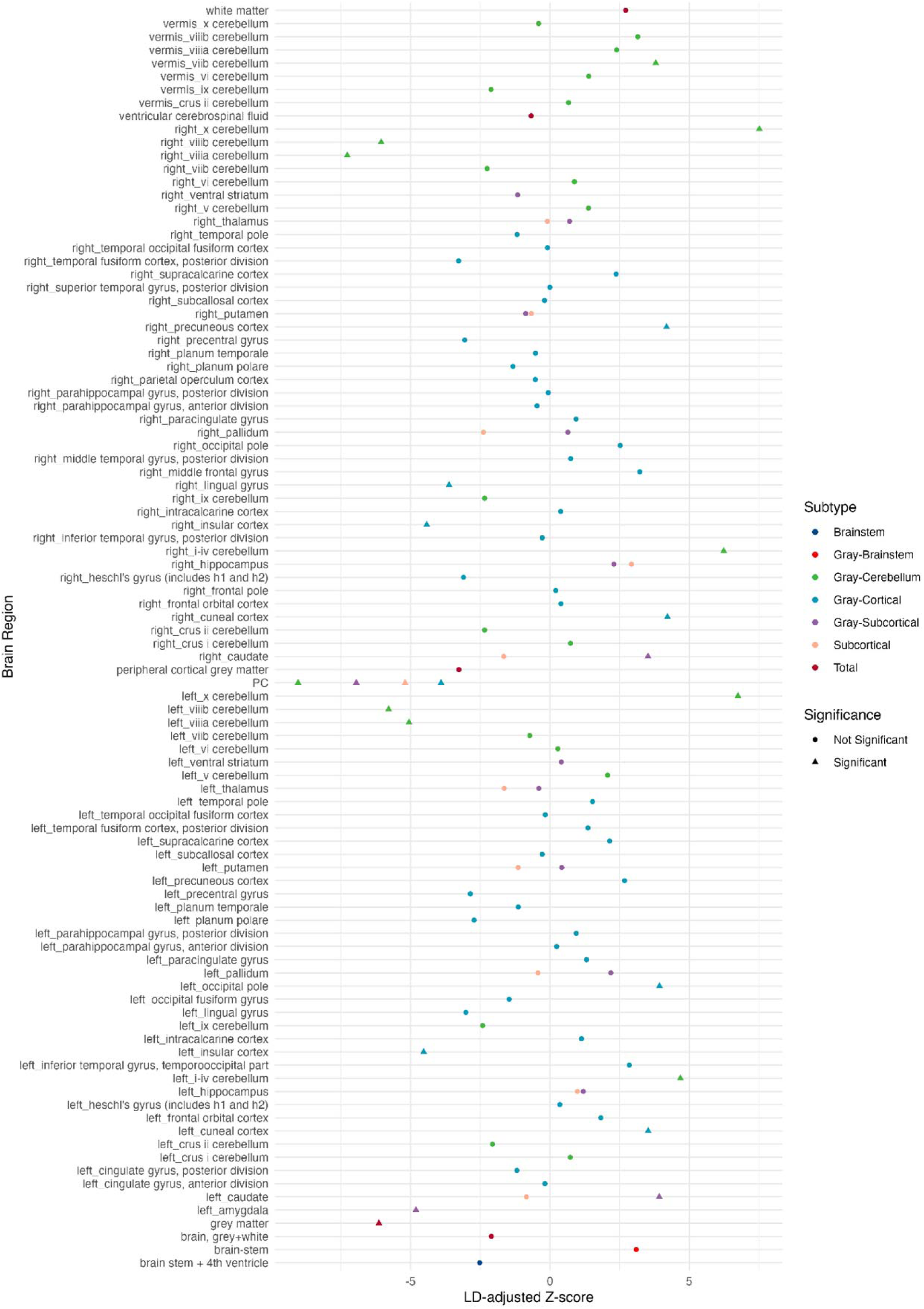
Structural features associated with BMI risk. LD-adjusted Z-scores are provided for region-specific associations of structural IDPs with BMI risk.

**Figure S9.**
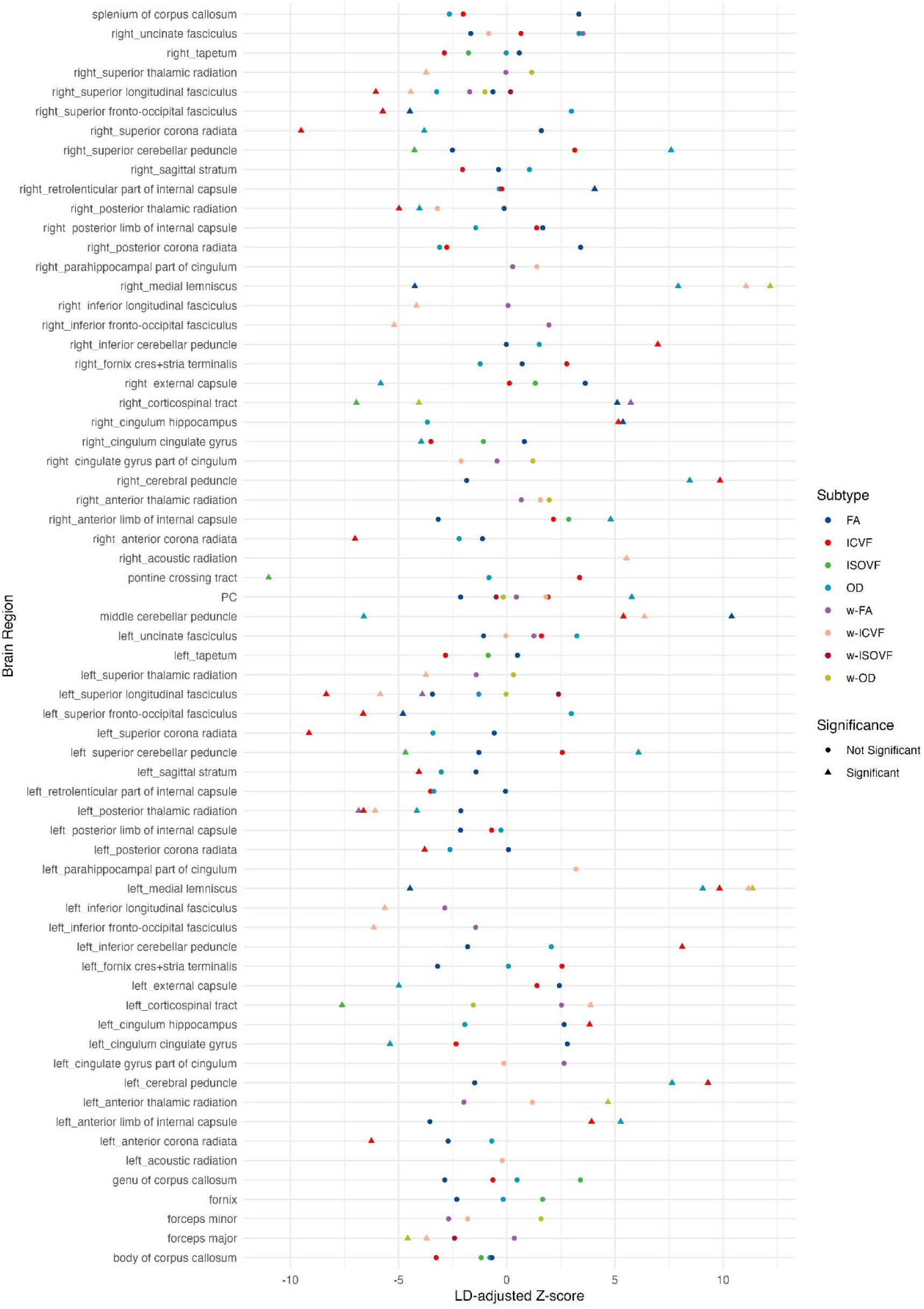
Diffusion features associated with BMI risk. LD-adjusted Z-scores are provided for region-specific associations of diffusion MRI IDPs with BMI risk.

## Notes

### Author Declarations

The Uniformed Services Universitys Human Research Protections Program Office and determined the project to be considered research not involving human subjects per 32 CFR 219.102(e)(1), and applicable DoD policy guidance. As such, this protocol does not require Institutional Review Board (IRB) review.

### Summary of Updates

Updated with several modifications in response to reviewers.

